# CUOREMA: Immersive Bio & Behavioral Feedback and Digital Interventions for Cardiac Rehabilitation – Exploratory Analysis

**DOI:** 10.64898/2026.05.15.26353188

**Authors:** Radoslava Švihrová, Davide Marzorati, Tania Odello, Giuliana Monachino, Tanya Staletti, Rob Tieben, René Luigies, Nicolette Bodewes, Werner Rutten, Glynn Barrett, Amber Bhogal, Tom Wilkinson, Athina Tzovara, Francesca Dalia Faraci

## Abstract

**Aims.:** Long-term adherence to cardiac rehabilitation (CR) remains low despite its proven benefits for secondary prevention. We explore the usability and engagement patterns of CUOREMA, a personalized mobile-health system integrating self-monitoring, wearable data, virtual coaching, and reinforcement learning-enhanced adaptive interventions to support lifestyle change during and after outpatient CR.

**Methods.:** In a six-month two-center observational study (N = 53, Switzerland and France), patients in outpatient CR used the CUOREMA app. Usability was assessed via validated and solution-specific questionnaires. Engagement patterns were identified using k-means clustering on weekly app usage. Clusters were compared by demographics and psychological themes of personal goals. Health-related questionnaires and the 6-minute walk test (6MWT) were collected at baseline and follow-up visits.

**Results.:** Three engagement clusters (dropout, sporadic, consistent) were identified and supported by matching patterns across app components and smartwatch wearing time. Clusters were not associated with demographic factors. Psychological themes of personal goals suggested that intrinsic motivation was more common among sustained users, while extrinsic motivation was more frequent among early dropouts. User experience was rated positively. One center showed significant 6MWT improvement, though the study was not powered for clinical outcomes. However, attrition was high: only 19% of participants used the app on more than 100 days, and questionnaire response rates declined from 55% at baseline to 13% at six months, and health-related drop-out can not be ruled out.

**Conclusion.:** Engagement variability driven by motivational rather than demographic factors is a central challenge in digital CR. Tailoring interventions to individual motivational profiles may improve adherence.

**Graphical Abstract:** 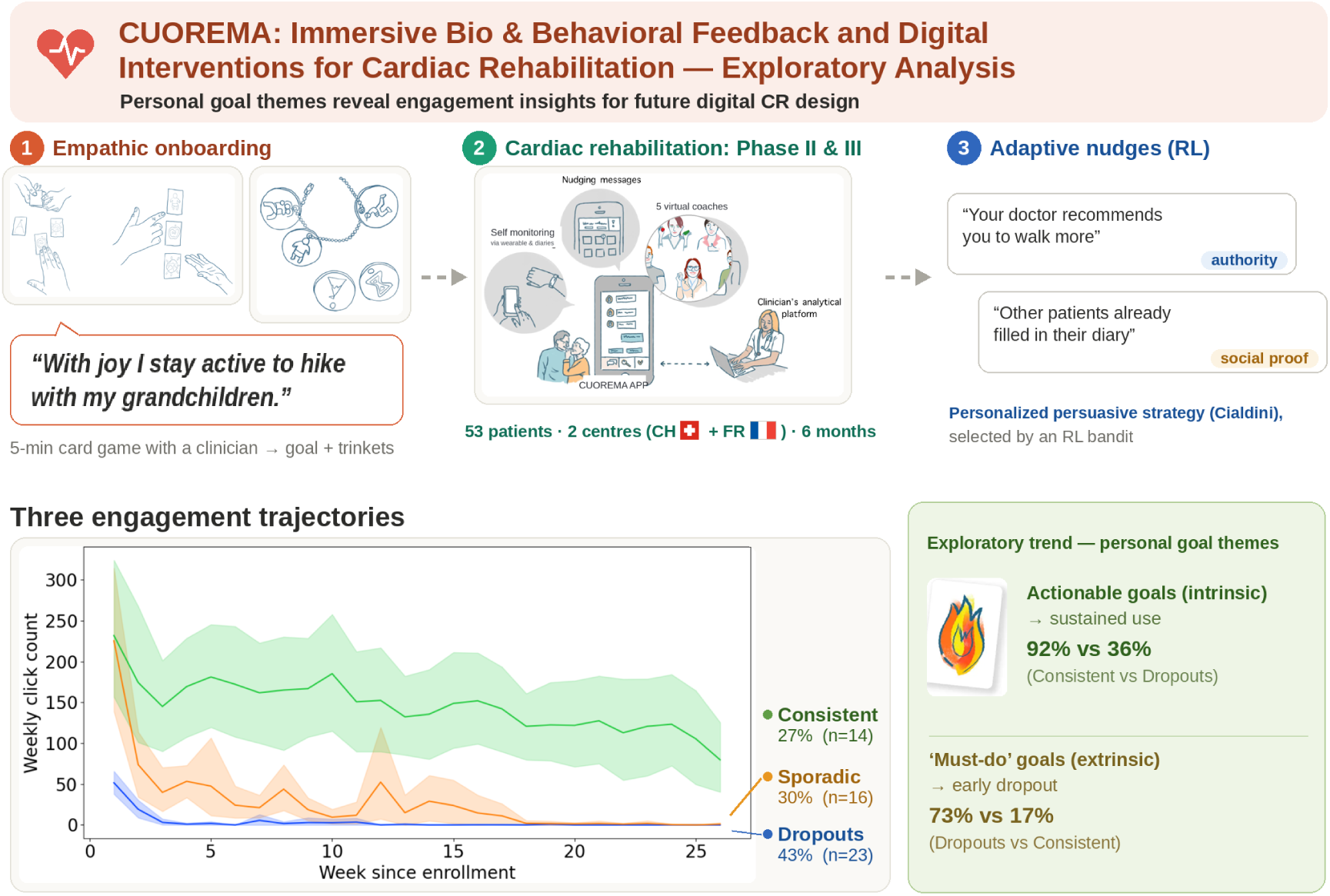

## 1 Introduction

Cardiovascular diseases can largely be prevented by improving health-related behaviors. The main lifestyle risk factors include physical inactivity, an unhealthy diet, tobacco use, and excessive alcohol consumption [43; 3]. The European Society of Cardiology (ESC) guidelines on cardiovascular disease prevention [68] highly recommend cardiac rehabilitation (CR) programs for individuals with multiple cardiovascular risk factors or following an acute cardiovascular event. CR programs aim to reduce the risk of recurrent cardiac events and improve quality of life through physical activity, heart-healthy nutrition, smoking cessation, stress management, and medication adherence [40]. Addressing these multiple lifestyle factors simultaneously requires sustained behavioral change, which remains a central challenge in CR.

CR programs are usually structured into three phases [65]: Phase I (early mobilization after an acute event), Phase II (comprehensive structured outpatient or inpatient rehabilitation, typically lasting around three months), and Phase III (long-term self-management). Despite the demonstrated effectiveness of outpatient CR [5], participation remains suboptimal. Many patients never enroll, and substantial numbers fail to complete the program. Barriers include older age, low socioeconomic status, limited health literacy, logistical challenges such as transportation or scheduling [20; 52], lack of social support [45; 70], and limited availability of rehabilitation centers and geographic distance discussed in the EUROASPIRE IV study [31]. Since failing to complete CR is associated with significantly worse cardiovascular outcomes [62], scalable, accessible, and personalized strategies to support long-term lifestyle change are needed.

Digital CR programs have demonstrated non-inferiority to conventional rehabilitation and potential for cost reduction [36; 64; 7]. Cardiac telerehabilitation was recently defined by the ESC as a quality indicator for CR centers [56], and the 2024 ESC guidelines on chronic coronary syndromes [69] include mHealth interventions as a Class I recommendation for improving adherence. However, studies to date have compared highly heterogeneous digital rehabilitation models, including telephone-based [46; 26], smartwatch-based [4; 38], and app-based monitoring [61; 21]. While several mHealth solutions aiming to cover multiple CR components have been developed, their adoption remains limited due to lack of reimbursement frameworks, regulatory heterogeneity, and insufficient evidence from large-scale trials.

Mobile health tools and wearable devices enable daily self-monitoring of behavior, physiological data, and symptoms [49; 37], and can deliver timely feedback and personalized motivation [16]. However, adherence to mHealth solutions is a central challenge: engagement often declines unless users perceive ongoing value or feel intrinsically motivated [71; 27; 54]. An effective mHealth lifestyle intervention should support the formation of long-term healthy habits, beginning with self-monitoring [6], continuing through repeated reinforcement, and ideally reaching automaticity [24; 23]. Timely and personalized digital health interventions have demonstrated effectiveness across various populations [74; 8; 35]. Incorporating social support through coaching or conversational interfaces can further enhance engagement and relatedness [59; 2].

In the “CUOREMA: Immersive Bio & Behavioral Feedback for a Better Heart Health" project (Eurostars grant E!115067), we developed a digital solution designed to support intrinsic motivation and sustained engagement for patients in phase II of the outpatient CR, using bio-behavioral feedback and gamification elements. The system integrates self-monitoring, educational content, virtual coaching, and personalized adaptive interventions delivered via push notifications. In the feasibility study we aimed to evaluate usability, characterize engagement patterns using data-driven methods, and explore associations between engagement and both demographic and psychological factors, similarly as done by [17], and to examine preliminary health-related outcomes.

## 2 Methods

### 2.1 System specification

The CUOREMA solution was designed with a lived experience approach, upon completion of interviews with the target population. It was not intended to replace the cardiologist and the CR team, but rather to complement them by providing a platform for simplified and comprehensive patient monitoring (e.g., physical activity, medication intake), guideline-based educational content aligned with ESC recommendations, and motivational support. Figure 1 shows the system schema. The final prototype consists of several components: mobile application, virtual companion, personalized adaptive interventions, and an analytical platform for medical professionals.

**Figure 1:**
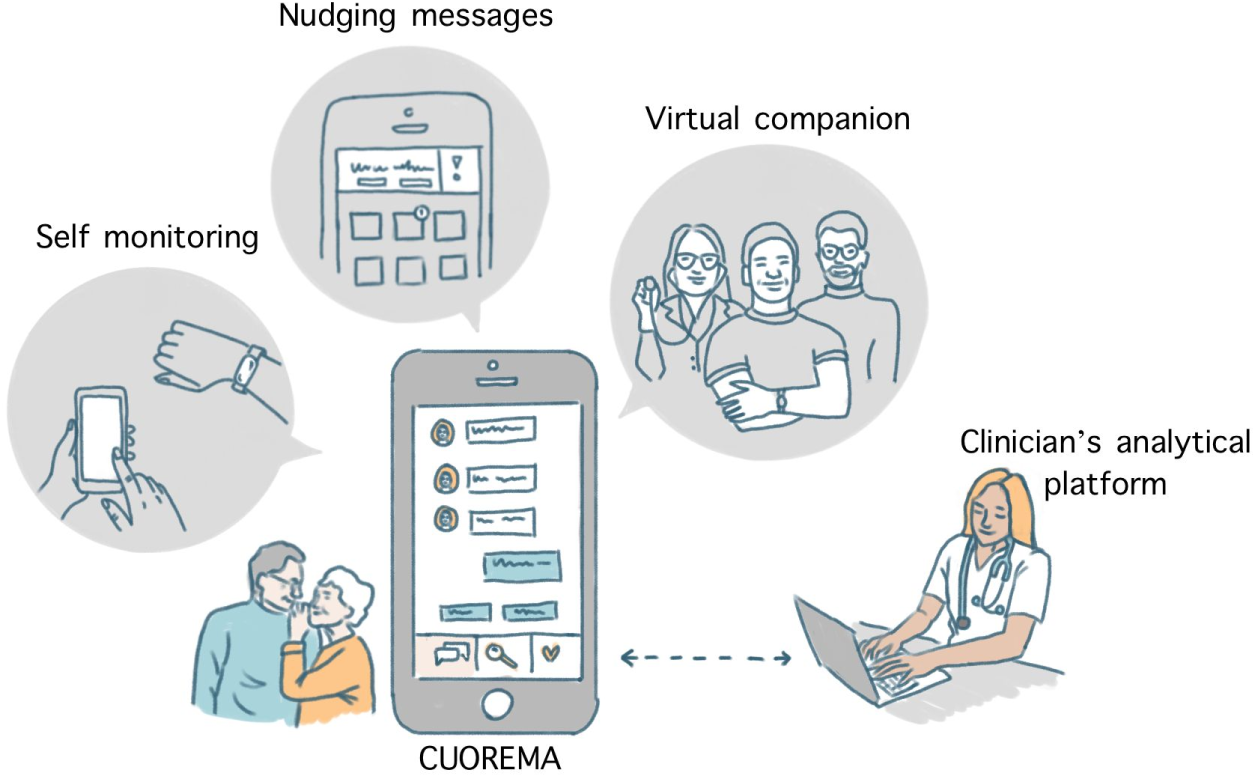
Components of the CUOREMA solution.

**Figure 2:**
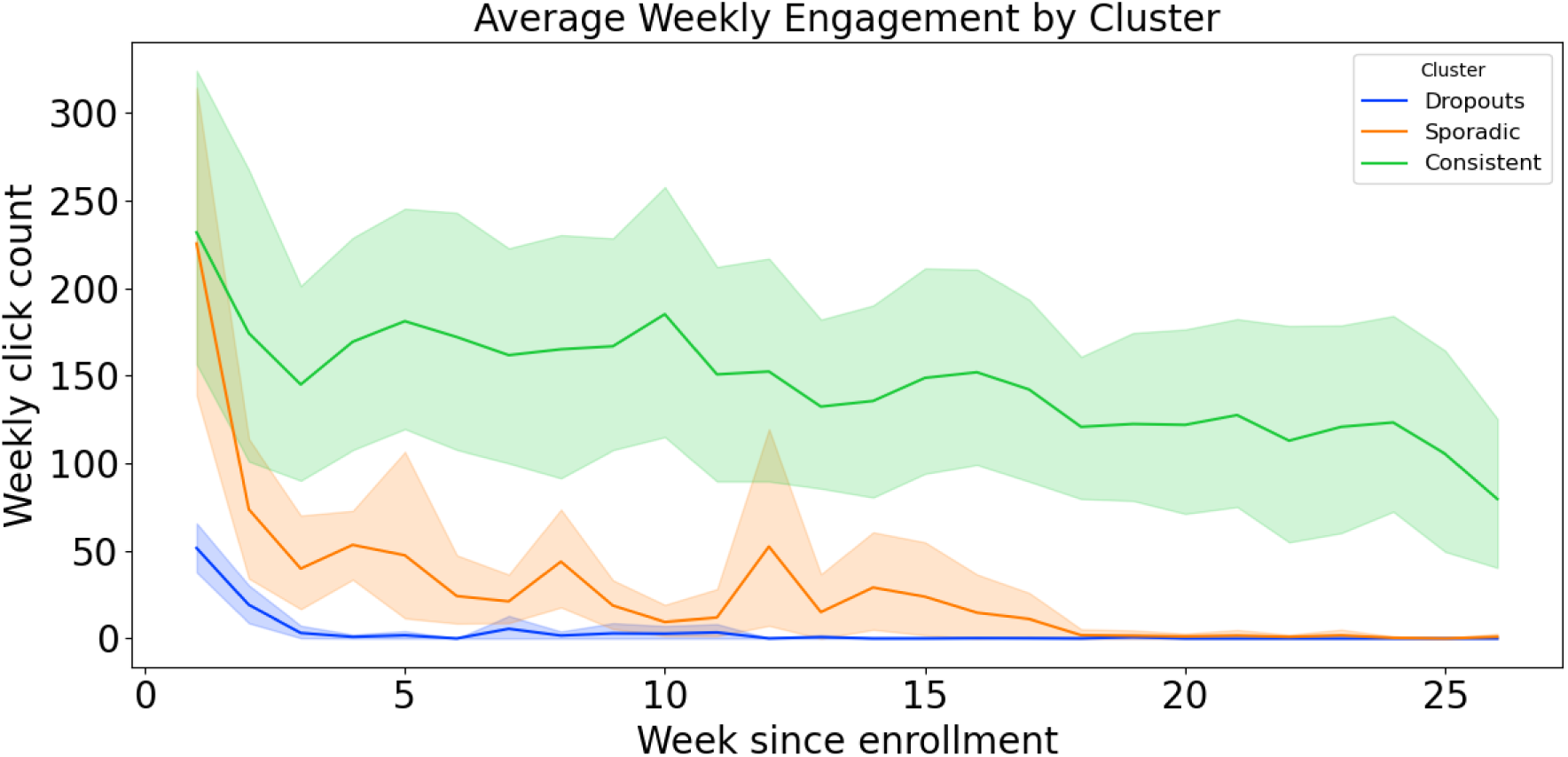
Average weekly click count in 3 engagement clusters with 95% confidence intervals.

**Figure 3:**
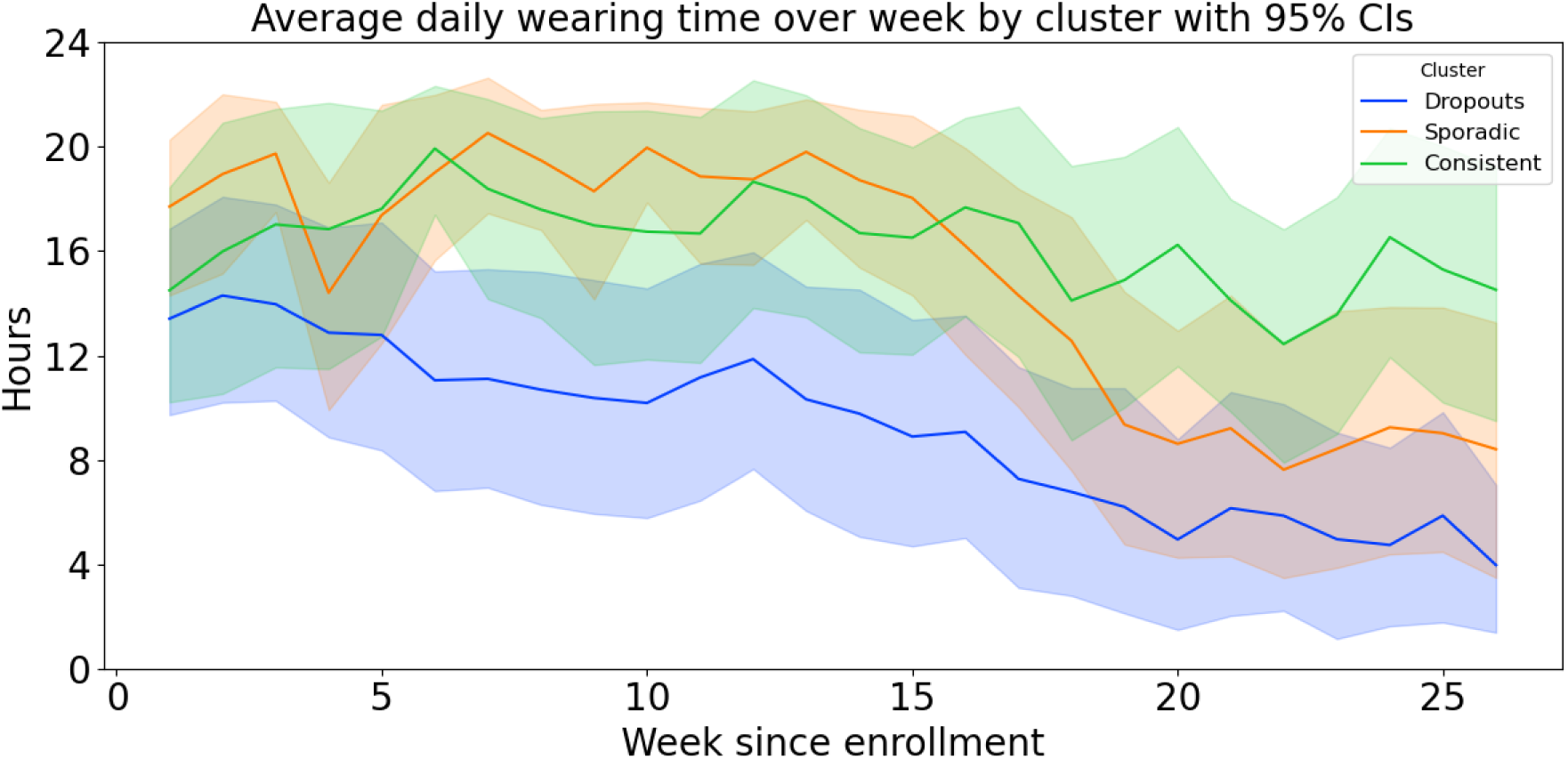
Average weekly wearing time across engagement clusters with 95% confidence intervals.

#### 2.1.1 Mobile application

CUOREMA was built as an extension of the MyHeart^1^ application (developed by my mhealth Limited), an NHS-certified platform tailored for CR patients. Key app sections include a *medication diary (MD)* for tracking medication adherence with optional push notification reminders, an *activity diary (AD)* for logging physical activities manually or automatically via wearable (including activity type, duration, and rate of perceived exertion on a 0–10 Borg scale [13]), *targets* for setting physiological goal ranges (steps, weight, BMI, blood pressure, glucose, cholesterol), and *educational and exercise videos* based on the patient’s diagnosis with a virtual reality component for tracking. For certain cardiovascular diagnoses (e.g. heart failure), symptom questionnaires (e.g. breathlessness, number of pillows used, fatigue, ankles swelling) appear daily at app launch. An analytical platform summarizes patient data for clinicians to support follow-up care. A detailed description of all app sections is provided in Supplementary Materials 5.1.

#### 2.1.2 Empathic onboarding and personal goals

Before first use, a health care professional (HCP) introduces the platform to the patient in a face-to-face conversation, explaining the purpose and use of the CUOREMA system. In addition, a five-minute physical card game is played to identify and verbalize the patient’s intrinsic motivations for recovery. Intrinsic motivation is important for psychological wellness and predicts an array of positive outcomes [55].

In the card game, patients are asked about the important things in their life, then pick semi-abstract illustrations that best represent these, eventually ending with three cards. The patient and HCP summarize this in a personal goal for motivation and recovery, illustrated in Figure 4 in Supplementary Materials 5.3. The goal and selected cards are entered in the app as part of onboarding. In addition, the patient receives three physical trinkets of the illustrations for a keychain (Figure 5 in Supplementary Materials 5.3).

**Figure 4:**
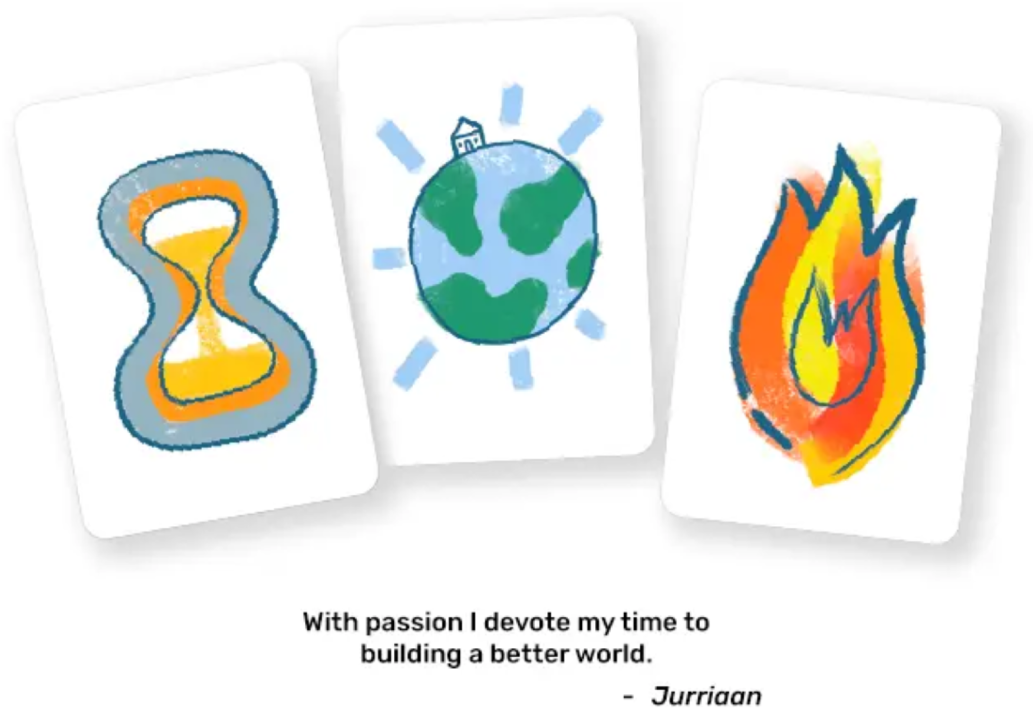
Example of selected cards and a personal goal formulated by the patient.

**Figure 5:**
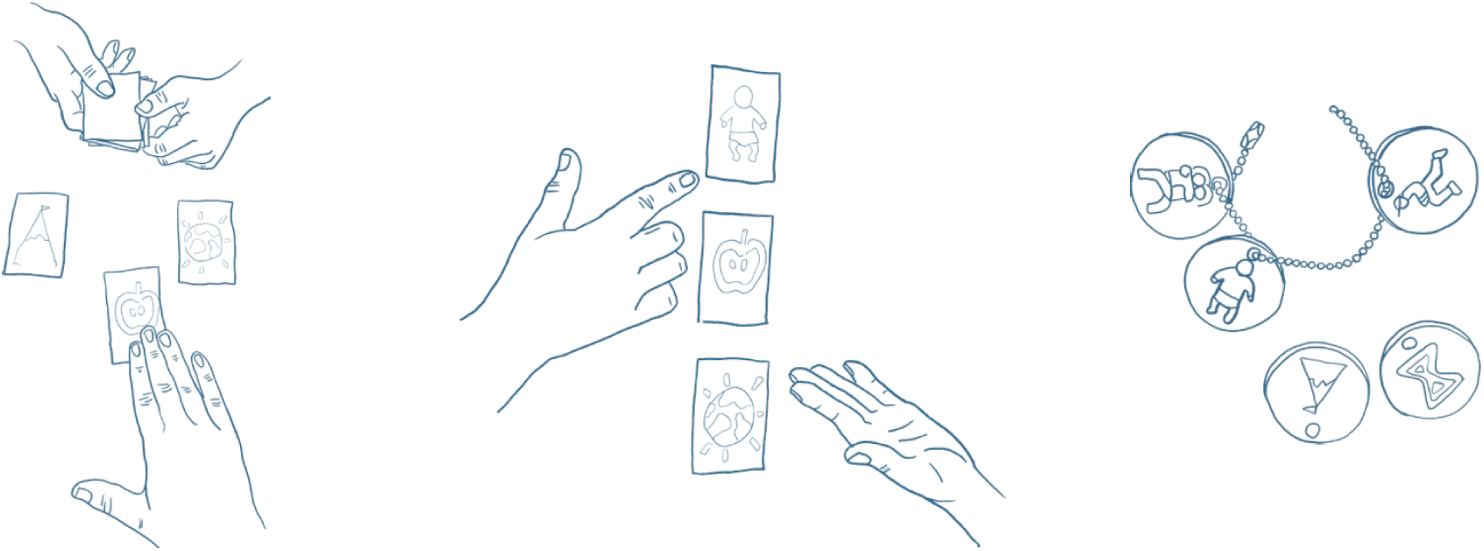
Illustration of physical card game for onboarding. Selected abstract cards help patient navigate to choose personal goal, and were provided to patient on a keychain.

#### 2.1.3 Virtual companion (VC)

Five virtual coaches (cardiologist, physiotherapist, dietitian, psychologist, smoking cessation coach) provide scripted personalized text interactions with the patient, giving recommendations, asking questions, and motivating the patient to adhere to the recovery guidelines. Each interaction starts with the personal goal and chosen illustrations to help recall and reinforce intrinsic motivation. Once a month, a digital review with the patient summarizes the progress and supports self-reflection and goal setting; the user receives a notification when the review is available. Illustrations and examples of virtual coaches conversations are presented in Figures 6, 7 and 8 in Supplementary Materials 5.3.

**Figure 6:**
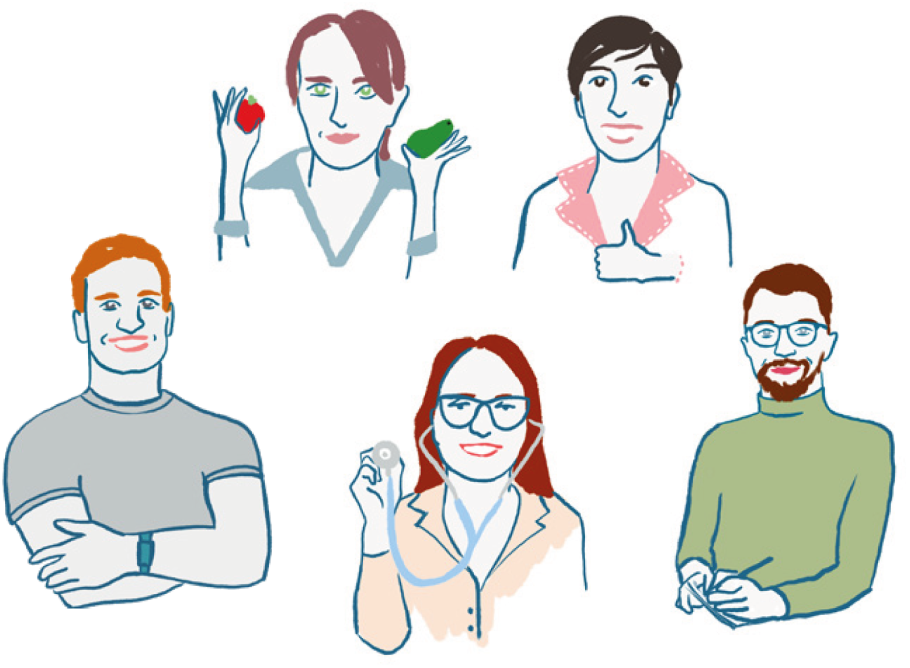
Illustration of 5 coaches in Virtual Companion.

**Figure 7:**
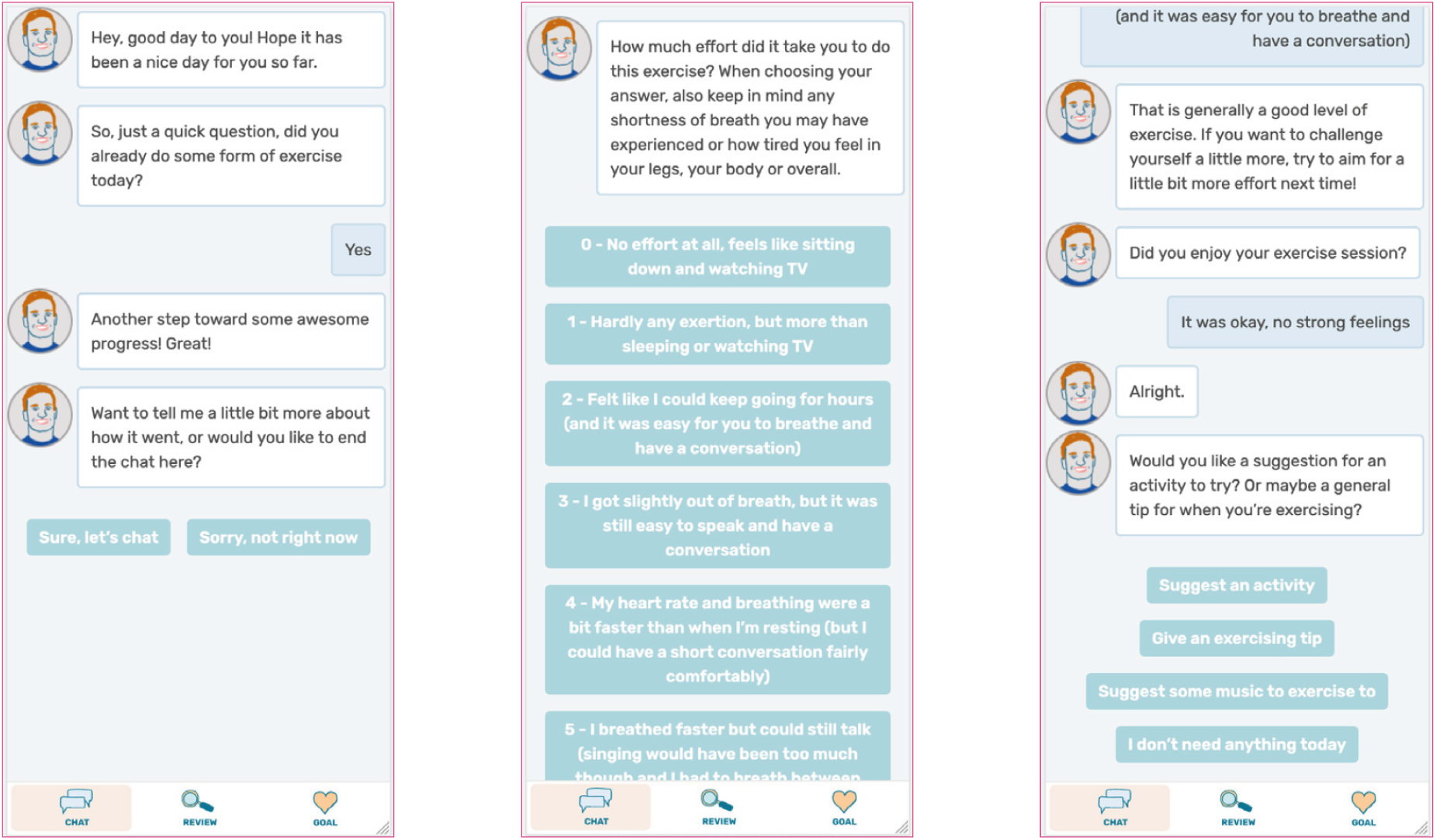
Example of conversation with Physiologist coach.

**Figure 8:**
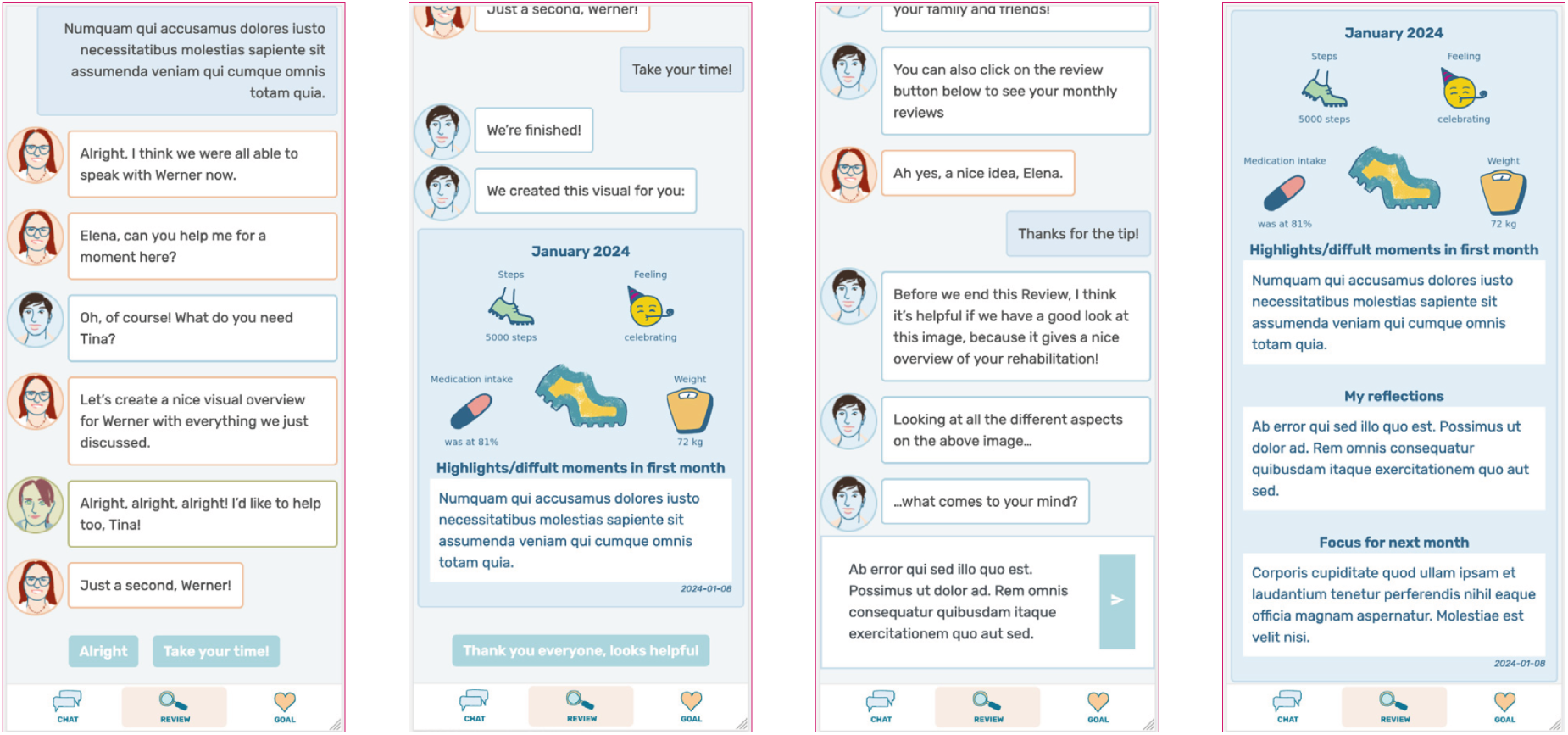
Example of monthly review in virtual companion.

#### 2.1.4 Personalized adaptive interventions

An adaptive digital intervention system tailors daily messages based on two data sources: (1) daily step count and activity data retrieved automatically from the wearable device, and (2) self-reported entries logged by the user into MD and AD. Since some medications are taken “as needed,” nudging for medication intake is limited to reminders to complete the MD. Weekly *step goal adjustment* prompts, asking the user to re-evaluate it with the physiotherapist, were sent each Monday at 10:00 if the goal had been met at least 10 times in the past 14 days. Upon availability of the monthly review in VC, a reminder to fill it in is sent on Friday and Saturday evening at 20:00, maximally three times. All messages were delivered as push or in-app notifications and upon clicking, the user was redirected to the respective app section.

##### Personalization of messages

All messages are clinician-approved, positively formulated and use gender-appropriate informal language. Some messages include user’s name (from introductory chat in VC), and some *bio-feedback* (e.g., current step count and target, or number of missed medications) to boost motivation. To enhance efficacy, all messages incorporate a *persuasive component* based on Cialdini’s six principles of influence [18], proven effective in digital health by Kaptein et al. [29]. For example, messages may leverage authority (“Your doctor recommends you to walk more”) or social proof (“Other patients in this program already filled in their diary”). Persuasive strategy selection uses a Bayesian multi-armed bandit (Thompson Sampling) [53] balancing exploration and exploitation, while building implicit user profiles. A sliding window accounts for changing user behavior over time [66]. With no prior user data, a uniform prior is assumed. Rewards are binary (success or failure), modeled with a Beta-Bernoulli distribution.

##### Daily timing and micro-randomization

Daily nudging messages are triggered daily at 17:00 based on MD and AD data. If all diary entries are complete and the daily step goal is met, a praise message is sent; otherwise, a nudging message is triggered [51; 63]. A follow-up check at 20:00 sends praise if the user subsequently met the target. If no step goal is set, a default of 5,000 steps is used. *Micro-randomization* [30] was applied to reduce user burden and enable causal inference on intervention efficacy, with a 20% probability of a control day (no intervention). This algorithm is detailed in [63] and summarized in Supplementary Materials 5.4.

### 2.2 Feasibility study

We conducted an observational two-center study to assess the feasibility of the developed solution, with the primary goal of gathering feedback on the system’s design and usability. Three study outcomes were defined: 1) evaluation of user experience, 2) analysis of patient interactions with app functions, and 3) analysis of health-related outcomes.

The study adhered to the Helsinki guidelines [73] and was approved by SwissEthics (BASEC ID: 2023-01413) and the Comité de protection des personnes Sud-Méditerranée I (23.04667.000297, 7601-000979-60). Data were pseudonymised and stored on secure servers in Europe and Switzerland. Access to the data was restricted to authorized study staff, and processing followed GDPR and local research ethics approvals. Participants had the right to withdraw; data collected before withdrawal were retained.

#### 2.2.1 Protocol and recruitment

The study was conducted over one year (01.03.2024 – 28.02.2025), with each participant enrolled for six months. Adults (*≥*18 years) in outpatient Phase II/III CR who owned a mobile device, were willing to use digital technologies, and had no unstable cardiovascular disease or disabling motor limitations were eligible. Full inclusion criteria are in Supplementary Materials 5.2.

Suitable participants were approached during the first month of outpatient Phase II CR. At onboarding (T0), HCPs provided complete information about the study, and interested participants signed informed consent. Demographic and clinical data were collected, and participants received an account in the mobile application to access all CUOREMA features and a Fitbit Charge 5 device, with instructions to use the Fitbit application only for synchronization. After setup, they played the physical card game, defined personal goals, and completed baseline questionnaires. After the first month, participants were asked to complete the user experience questionnaire. The first follow-up visit (T1) was scheduled after three months, corresponding to the end of Phase II and the start of Phase III. The second follow-up visit (T2) was scheduled after another three months, i.e., six months from onboarding. At both follow-up visits, clinical measurements were repeated and questionnaires administered. At T2, the Fitbit device was returned. All questionnaires were distributed digitally as a chat within the Virtual Companion and remained open for four weeks; if not completed, automated reminders were sent and HCPs were instructed to contact participants via phone. The data sources and the measures derived from them are described in the following section. Adverse events were monitored continuously throughout the study and recorded by the responsible HCPs, both at follow-up visits and through spontaneous participant reports. The data sources and the measures derived from them are described in the following section. All study components were available in English, Italian, French, and German. The protocol is summarized in Figure 9 in Supplementary Materials.

**Figure 9:**
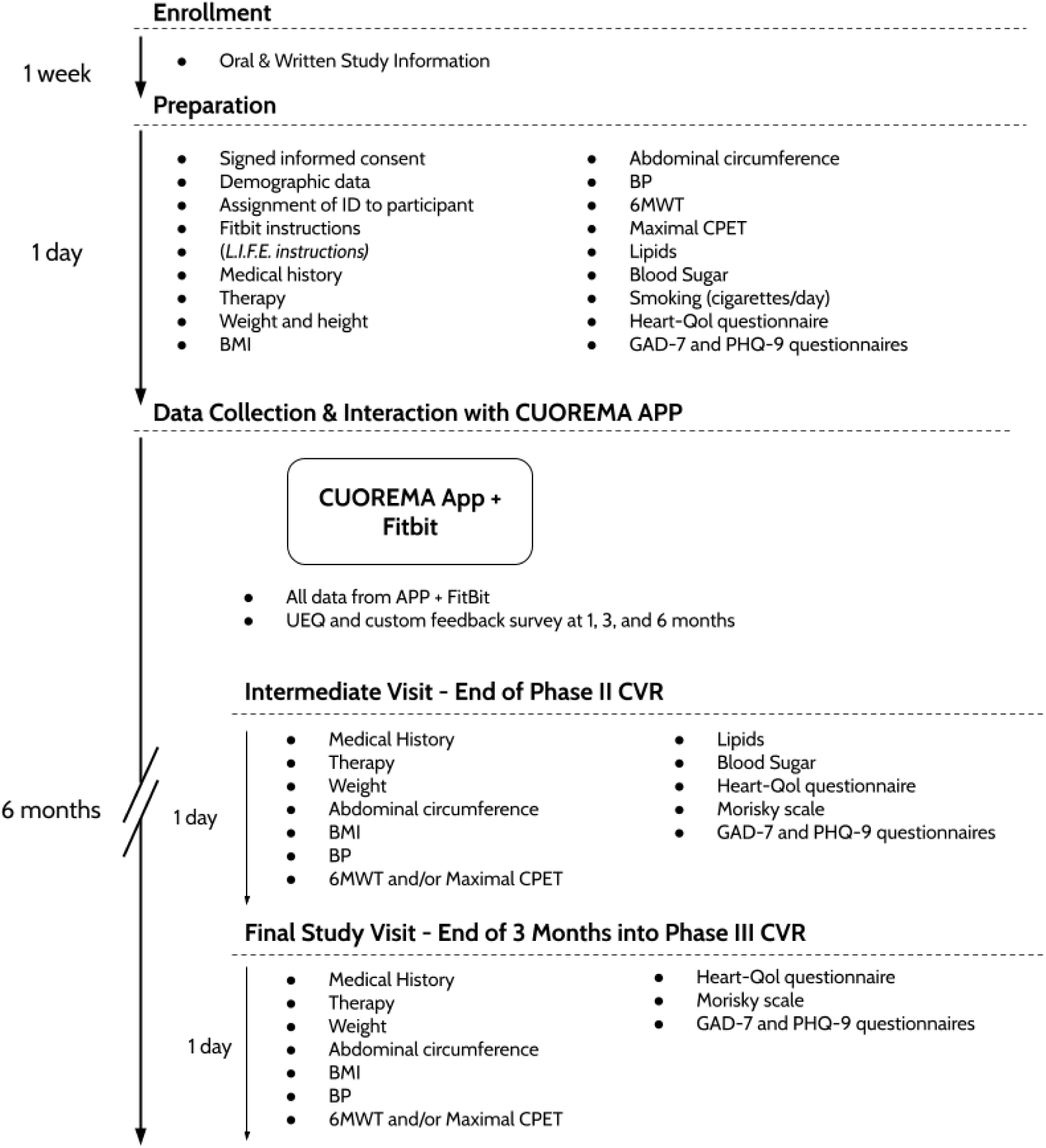
Schema of the study protocol for the CUOREMA feasibility study.

#### 2.2.2 Study outcomes and data sources

All data were collected within the protocol described above and pseudonymised before analysis. Demographic information (age, sex, occupation and work years, household composition, weight, BMI, and smoking habits) was recorded at onboarding (T0). Clinical information, comprising cardiological history, medication therapy, vital parameters (blood pressure, heart rate from ECG, abdominal circumference, blood sugar, cholesterol, and triglycerides), and clinical tests (the 6-minute walking test (6MWT), treadmill stress test, and cardiopulmonary exercise testing [48; 68; 9]), was collected at T0, T1, and T2. Interactions with virtual companion were logged, personal goals were translated into English and assigned psychological themes by a social science expert. Sending of notifications time was stored, as information about receiving them was unavailable.

For each outcome we specify the data source and the corresponding *quantifiable measures*:

1. **Evaluation of user experience.** Assessed through the 26-item User Experience Questionnaire (UEQ) [58], covering attractiveness, pragmatic quality, and hedonic quality, and solution-specific questions, all collected after the first month and at T1 and T2:

(a) Results and temporal changes in the UEQ.
(b) Results and temporal changes in solution-specific questions on the perceived efficacy and utility of individual CUOREMA components.
2. **Analysis of patient interactions with app functions.** Derived from application logs (login timestamps and background interaction events), digital diary entries with timestamps, virtual coaches interactions, and wearable data extracted after the study end via the Fitbit API:

(a) Login sessions and engagement.
(b) Time to dropout (last opening of the app).
(c) App components usage (targets, activity diary, medication diary, virtual coaches).
(d) Wearable adherence (daily wearing time).
3. **Analysis of health-related outcomes.** Based on clinically validated questionnaires administered at T0, T1, and T2, namely Health-related Quality of Life (Heart-QoL) [44], the 7-item Generalized Anxiety Disorder scale (GAD-7) [60], the 9-item Patient Health Questionnaire (PHQ-9) [32], and the 8-item Morisky scale [11; 15], together with the 6-minute walking test (6MWT) collected at the same visits:

(a) Results and temporal changes in the validated questionnaires.
(b) Change in 6MWT performance throughout the study.

Demographic variables (age, sex, and rehabilitation center) were used as grouping factors for between-group comparisons of the study outcomes, which were complemented by paired within-subject tests to assess change over time.

### 2.3 Statistical methods

**Continuous and ordinal outcomes** were compared using the Wilcoxon rank-sum test and Mann–Whitney U test [25]; for more than two groups, the Kruskal–Wallis H-test [33] was used. Pearson correlation was used for associations [10].

**Proportions in contingency tables** were tested based on the data generating process: Fisher’s exact test [22] for hypergeometric data (both frequencies in rows and columns known), Boschloo’s test [14] for binomial (frequency in rows or columns known), and *χ*^2^-test [1] for multinomial distributions. Bonferroni correction controlled the family-wise error rate in multiple hypothesis testing [12]. Where appropriate, (generalized) regression models were employed [42]. **Time-to-event analysis** used a Cox proportional hazards (Cox-PH) model [57] with time-stable covariates (age, sex, center) to avoid violations of the PH assumption [28].

To **group** participants with similar engagement patterns, k-means clustering [34] was applied to binary weekly engagement indicators (1 if app opened at least once per week, 0 otherwise). The optimal number of clusters was determined by the elbow method.

Analyses used Python packages scipy [67], lifelines [19], and scikit-learn [47].

## 3 Results

A total of N=53 patients were recruited from OSCAM (Ospedale Malcantonese, Switzerland) and Corbie (Centre Hospitalier de Corbie, France). Patients were enrolled on a rolling basis (first: 26.02.2024, last: 04.12.2024); 18 (34%) patients (2 from OSCAM, 16 from Corbie) enrolled after 01.09.2024, preventing them from completing the full six months. Demographics are summarized in Table 1.

**Table 1:** Demographic information. Continuous variables are reported as median [*p*_25_*, p*_75_], and categorical variables as *N* (%).

|  |  | Overall | Corbie (FR) | OSCAM (CH) | p-value |
| --- | --- | --- | --- | --- | --- |
| Participants |  | 53 | 35 | 18 |  |
| Age |  | 56.0 [51.0,64.0] | 55.0 [50.5,61.5] | 60.5 [53.2,65.5] | 0.15 |
| BMI | T0 | 28.3 [23.5,32.6] | 29.6 [23.6,34.1] | 26.0 [23.5,31.7] | 0.341 |
|  | T1 | 25.8 [24.3,29.3] | – | 25.8 [24.3,29.3] | N.A. |
|  | T2 | 25.6 [23.9,31.5] | – | 25.6 [23.9,31.5] | N.A. |
| Work hours per week |  | 35.0 [35.0,40.0] | 35.0 [35.0,38.8] | 40.0 [25.5,40.5] | 0.617 |
| Rest Heart Rate - T0 |  | 66.0 [59.0,78.0] | 63.0 [57.0,72.5] | 71.0 [62.8,85.5] | 0.031 |
| Sex | F | 14 (26%) | 8 (23%) | 6 (33%) | 0.624 |
| Smoker | Yes | 11 (21%) | 4 (11%) | 7 (39%) | 0.039 |
|  | Ex | 17 (32%) | 11 (31%) | 6 (33%) |  |
|  | Pack years | 19.4 [14.7,30.5] | 20.0 [15.0,31.0] | 15.0 [4.2,30.0] | 0.255 |
| Comorbidities | Hypertension | 26 (49%) | 16 (46%) | 10 (56%) | 0.698 |
|  | Ischemia | 32 (60%) | 30 (86%) | 2 (11%) | < 0.001 |
|  | Arrhythmias | 7 (13%) | 7 (20%) | 0 (0%) | 0.108 |
|  | Diabetes | 9 (17%) | 7 (20%) | 2 (11%) | 0.667 |
|  | Cholesterolemia | 44 (83%) | 30 (86%) | 14 (78%) | 0.732 |
|  | Cardiac Interventions | 33 (62%) | 32 (91%) | 1 (6%) | < 0.001 |

**Table 2:**
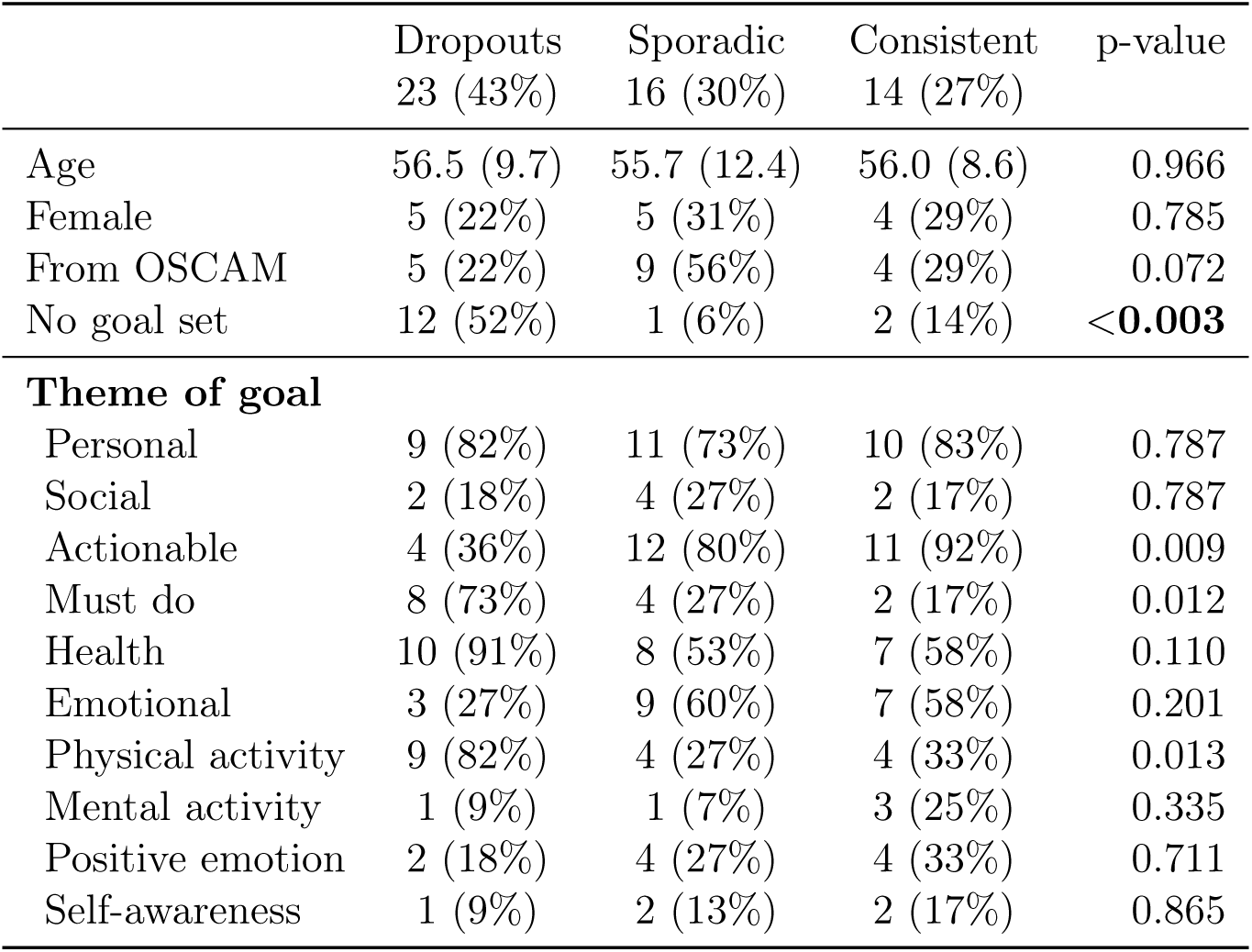
Participant characteristics by engagement cluster. Age: mean (sd), Kruskal-Wallis H-test. Others: N(%), *χ*^2^-test.

### 3.1 Personal goals

Of 53 participants, 52 (98%) stored selected cards in the VC, of them 38 (73%) set a personal goal, and never updated it. Only one of the 15 non-selectors was from OSCAM (Boschloo’s test, *p* = 0.006). No significant differences in age (Mann-Whitney U, *p* = 0.521) or sex (*χ*^2^, *p* = 0.089) were found between goal selectors and non-selectors after Bonferroni correction (*α* = 0.017).

A social science expert tagged the formulated goals with one or more of 10 psychological themes:

1. **Personal goal**: Individual focused goal
2. **Social goal**: Social/relatedness focused goal
3. **Actionable goal**: Concrete actionable goal
4. **Must do goal**: Compliance focused goal
5. **Health goal**: Health/Medical focused goal
6. **Emotional goal**: Emotional/personal (non-medical) goal
7. **Physical activity goal**: Activity/physical focused goal
8. **Mental activity goal**: Interest/mental/hobby focused goal
9. **Positive emotion goal**: Happiness/positive emotion focused goal
10. **Self-awareness goal**: Awareness/personal development focused goal

Goal themes were compared by center, age, and sex among goal-setting participants. After Bonferroni correction (*α* = 0.005), no significant differences emerged. Full results are in Supple-mentary Table 4.

**Table 3:**
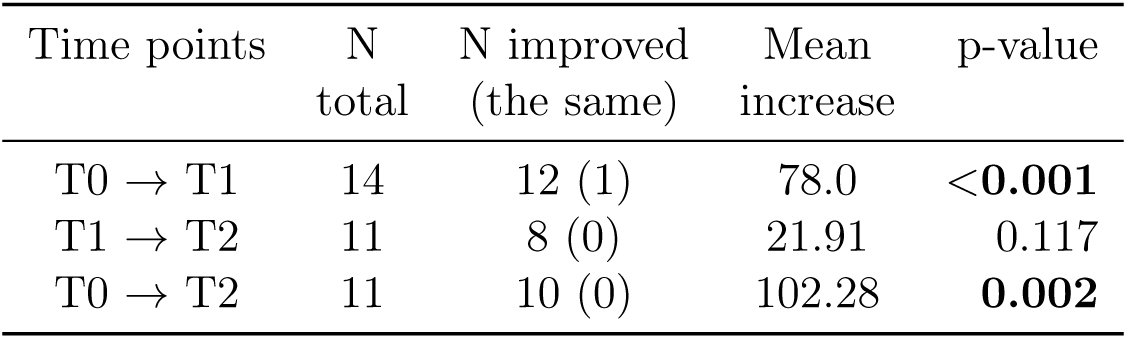
One-sided paired Wilcoxon test for improvement in 6MWT (OSCAM).

**Table 4:** Summary of goal themes, reporting user *counts* (p-values) for differences in character-istics. Statistical tests used Bonferroni corrected *α* = 0.005.

| Theme of goal | N (%) | OSCAM | F | Age |
| --- | --- | --- | --- | --- |
|  | Tot=38 | Tot=17 | Tot=13 |  |
| Personal | 30 (79%) | 10 (0.019) | 11 (0.843) | 0.680 |
| Social | 8 (21%) | 7 (0.019) | 2 (0.843) | 0.680 |
| Actionable | 27 (71%) | 14 (0.307) | 8 (0.579) | 0.759 |
| Must do | 14 (37%) | 4 (0.233) | 5 (1.0) | 0.856 |
| Health | 25 (66%) | 8 (0.065) | 9 (1.0) | 0.414 |
| Emotional | 19 (50%) | 13 (0.009) | 6 (1.0) | 0.804 |
| Physical activity | 17 (45%) | 6 (0.468) | 6 (1.0) | 0.402 |
| Mental activity | 5 (13%) | 5 (0.029) | 2 (1.0) | 0.778 |
| Positive emotion | 10 (26%) | 7 (0.133) | 3 (1.0) | 0.740 |
| Self awareness | 5 (13%) | 2 (1.0) | 1 (0.831) | 0.846 |

### 3.2 Evaluation of user experience

#### Results and temporal changes in the UEQ

UEQ responses were collected from 9, 14, and 7 participants at the first month, T1, and T2, respectively. Of 20 total respondents, one completed two consecutive questionnaires and four completed all three, while the rest provided a single response. All median values were positive (Supplementary Figure 10). The median perspicuity, reflecting ease of learning, appeared to increase over time, suggesting growing familiarity with the system, while novelty and attractiveness tended to decrease.

#### Solution-specific questions

Solution-specific questions (7-point Likert scale) were completed by 9, 13, and 7 participants at the three time points. Initial responses were mostly positive, while later responses tended toward neutral. Most participants initially felt the program supported their CR goals, physical activity, vital sign monitoring, and health awareness, though these effects declined at the second assessment. The medication diary reminders were consistently rated negatively. Motivational messages were rated as engaging early on but received mixed later feedback, and virtual companion reported use dropped over time. No clear worsening trends were observed in negative responses. Detailed positive and negative response rates are in Supplementary Materials 5.7.

### 3.3 Analysis of patient interactions with app functions

#### 3.3.1 Login sessions and engagement

All 53 participants logged in at least once. Engagement varied: 5 (9%) logged in only once, 17 (32%) used the app on *≤*10 days, and 10 (19%) engaged on *≥*100 days. A beta regression of the proportion of active days (days with at least one interaction divided by total study days) with CR center, age, and sex as predictors showed no significant effects (*p >* 0.5), indicating demographics did not influence app adherence. Weekly trends revealed strong early engagement: 44 participants (83%) reached peak use within four weeks, and 37 (70%) in the first week, suggesting high initial motivation followed by a gradual decline over time. Of the 18 late enrollees, 11 were classified in the Dropout cluster; however, their engagement patterns were consistent with the cluster definitions rather than an artifact of shorter observation time.

##### Engagement clusters

K-means clustering was applied to binary weekly engagement indicators to identify usage patterns. Using the elbow method, three clusters were selected as optimal. These represented distinct trajectories (Figure 2): (1) **Dropouts** (N=23, 43%) – high initial use followed by rapid decline, (2) **Sporadic** (N=16, 30%) – low overall use with a peak around the medical visit at T1, and (3) **Consistent** (N=14, 27%) – steady usage throughout the study period.

As shown in Table 2, no significant differences in age (Kruskal-Wallis), sex, or CR center (*χ*^2^ tests) were found across clusters, suggesting these demographic factors did not influence engagement. After Bonferroni correction (*α* = 0.003), a borderline significant difference emerged in goal-setting, with the highest proportion of non-set goals in the Dropout cluster (52%).

Among participants who did set goals, differences in goal themes, although not statistically significant after Bonferroni correction, showed interesting trends: the actionable theme (intrinsic motivation) was prevalent for Consistent and Sporadic participants. Conversely, the must do and physical activity themes (compliance-driven, extrinsic motivations) were most prevalent in Dropouts.

#### 3.3.2 Time to dropout

To assess the time to dropout, an event was defined as a participant’s first day of non-use following their last app interaction during the 182 days (six months) in the study. Among 35 participants enrolled before 01.09.2024, 15 experienced a dropout event (43% dropout rate), including 3 who disengaged after the first day. Of these 15 events, 11 (48%) occurred in the Dropout cluster, 3 (19%) in Sporadic, and 1 (7%) in Consistent. Some members of the Dropout and Sporadic clusters did not experience an event due to right-censoring or minimal re-engagement near the study’s end. A *χ*^2^-test rejected independence between clusters and dropout events (*p* = 0.017), supporting cluster validity. A Cox-PH model fitted using a Breslow estimate of the baseline hazard revealed no significant difference in hazard ratios between centers (*α* = 0.05), consistent with previous findings. Reported drop-out reasons included work commitments (3), discomfort with being monitored (1), new cardiac events (1), orthopedic problems (1), cancer diagnosis (1), and personal reasons (1), and several participants disengaged without explanation.

#### 3.3.3 App component usage

##### Targets

In total, 7 participants set at least one goal in the targets section (2 from OSCAM) most commonly for steps (3) and weight (4). Only 1 participant adjusted their weight goal, each three months. By engagement cluster, 2 were Dropouts, 3 Sporadic, and 2 Consistent.

##### Activity diary

Of 53 participants, 51 (96%) connected the provided Fitbit wearable for passive monitoring, with 50 having at least one automatically detected activity. *Manual activity entries* were made by 18 participants, of whom 8 recorded activities more than five times, and 15 participants manually entered daily step counts. Manual and automatic activity counts showed moderate positive correlation (*ρ* = 0.499, *p* = 0.0002). No significant differences were observed between centers in manual entries (*χ*^2^, *p* = 0.139), or age between manual entry users and non-users (Mann–Whitney U, *p* = 0.233). Regression of manual entries on personal goal clusters revealed no significant effects.

##### Medication diary

Medication therapy was created by 36 (68%) participants, medication diary by 21 (40%), and 9 (17%) participants activated medication reminder. At the corrected significance level (*α* = 0.017), only the difference between medication therapy creation between centers was statistically significant(Boschloo’s test, *p* = 0.002), as summarized in Supplementary Table 5. Exploration of entry patterns revealed that some participants entered most medication intakes at once, suggesting retrospective input rather than daily self-monitoring. A *χ*^2^-test rejected independence between engagement clusters and MD usage (*p <* 0.001): no Dropout participants used the MD, compared to 9 (56%) Sporadic and 12 (86%) Consistent. MD and AD usage were also significantly associated (*χ*^2^-test, *p* = 0.003), suggesting participants tended to either use both diaries or neither.

##### Virtual coaches

Of 17 (32%) participants who engaged with the virtual coaches, (29%) interacted with all of them. Users typically interacted only once, with a few returning sporadically; one highly engaged participant recorded over 70 interactions with the physiologist alone. Details on interaction counts are reported in Supplementary Table 6. The usage was significantly associated with engagement clusters (*χ*^2^-test, *p* = 0.006): 2 (9%) from Dropouts, 8 (50%) from Sporadic, and 7 (50%) from Consistent used the coaches. No significant differences in age, sex, or center were found between users and non-users. These results reinforce the lack of demographic effects on engagement and support cluster validity.

#### 3.3.4 Wearable adherence

Daily wearing time, estimated from Fitbit heart rate data at minute resolution from 50 participants, was aggregated into weekly averages, presented in Figure 3 by engagement clusters. Although 95% confidence intervals overlapped, trends in weekly wearing time aligned with the engagement clusters, further supporting their validity. Median [*p*_25_, *p*_75_] of weekly averages of daily wearing time was 18.7 (10.8; 21.6) hours for women and 13.5 (8.5; 18.0) for men, with no statistically significant difference (Mann-Whitney U, *p* = 0.093), but a trend toward higher adherence in women. No correlation between age and wearing time was found (Pearson’s *ρ* = 0.061, *p* = 0.673).

### 3.4 Analysis of health-related outcomes

#### 3.4.1 Validated questionnaires

At T0, T1, and T2, 28 (29 for Heart-QoL), 15, and 7 responses were collected, respectively. As only three participants completed all assessments and only one completed T1 and T2, inference across all time points was not feasible. A one-sided paired Wilcoxon test comparing T0 and T1 (Supplementary Table 13) found no significant effects after Bonferroni correction (*α* = 0.012), though trends suggested stability or potential improvement across measures. Baseline scores were already favorable: median GAD and PHQ scores indicated no or minimal anxiety and depression, and Morisky scores indicated good medication adherence (Supplementary Tables 15–18).

To assess differences between engagement clusters, questionnaire responses at T0 were compared between Sporadic (N=13) and Consistent (N=11) clusters; the Dropout cluster had too few responses (N=4). After Bonferroni correction (*α* = 0.008), no differences were significant, though trend in lower QoL Total score (*p* = 0.051) in the Consistent cluster was observed, warranting further investigation. More details are reported in the Supplementary Table 14.

#### 3.4.3 6-Minute Walk Test

At OSCAM, all 18 participants completed the 6MWT at T0, 14 at T1, and 11 at T2. In Corbie, only one participant was assessed, and only at T0. Median 6MWT distance improved from 527 m at T0 to 605 m at T1 and 603 m at T2 (full summary statistics in Supplementary Table 19). One-sided paired Wilcoxon tests (Table 3) on Bonferroni corrected *α* = 0.017 showed significant improvements between T0–T1 and T0–T2, but not between T1–T2, suggesting that the main improvement occurred during Phase II, with subsequent stabilization during Phase III.

#### 3.4.3 Adverse events

No serious adverse events were considered related to the usage of the CUOREMA solution. A few participants reported skin irritation from the smartwatch, which resolved after they were provided with an alternative wristband. Three participants withdrew following intercurrent medical events, namely one new cardiac event, one orthopedic condition, and one cancer diagnosis.

## 4 Discussion

We developed and evaluated CUOREMA, a gamified and adaptive mobile health solution for patients during and after outpatient CR. CUOREMA is a research prototype evaluated under ethics approval as part of this feasibility study. It is not a certified or marketed medical device. All findings reported here relate only to usability, feasibility and exploratory observations within this study. No conclusions on clinical efficacy can be drawn at this stage, and further research will be required before any regulatory evaluation.

During *unstructured interviews*, patients identified the AC and VC as the most used features, while the MD was least used. Some participants appreciated the goal-setting card game for maintaining motivation. Reported issues included a complex MD interface for everyday use, limited personalization (e.g., inability to customize the home page or choose conversation topics with the coach), a less engaging design compared to the Fitbit application, and one patient reported excessive notifications. Overall, patients expressed a preference for an even more *user-centered and adaptive design*. After the study, HCPs noted that at OSCAM, medication therapy was entered into the MD together with patients during onboarding, whereas in Corbie, patients managed it independently, which likely explains Corbie’s lower adoption of the MD but reflects a more realistic usage setting. While the unstructured feedback points to the need for improved personalization, the available dropout reasons were predominantly medical or practical in nature, although incomplete reporting limits broader conclusions about attrition.

The *exploratory statistical analysis* aimed to assess user experience and explore patient interaction with CUOREMA to inform future system development. Preliminary efficacy was also explored using validated health-related questionnaires and clinical tests at baseline and follow-up, although the study was not powered to detect clinical outcomes and results should be interpreted as exploratory. Analyses addressed three main study outcomes, yielding the following findings:

1. Patients and clinicians reported satisfactory *user experience* with no apparent worsening and emphasized the importance of future user-centered refinement.
2. Three *engagement clusters* were identified and validated through consistent usage patterns across app components, with no evidence of demographic influence. The presence of *psychological themes* suggested intrinsic motivation (actionable theme) in Consistent and Sporadic clusters, whereas Dropouts appeared more extrinsically motivated (physical activity and must do themes).
3. No deterioration was observed in *the results of validated questionnaires*. A statistically significant improvement in *walking performance* was observed at OSCAM, although clinical relevance was not assessed and selective dropout for health-related reasons cannot be ruled out.

Although absence of evidence does not imply evidence of absence, and despite the small sample size and high drop out, these results offer valuable insights into engagement patterns among CR patients. For further evaluation of temporal changes in engagement metrics across clusters, a larger and more diverse sample is needed to enhance generalizability.

From a *psychological perspective*, behavioral change unfolds through several stages [50] and, according to self-determination theory, is primarily driven by intrinsic rather than external motivation. In clinical settings, distinguishing passive compliance, i.e., following medical advice without personal commitment, from active adherence, where individuals take responsibility for their health, is essential [41]. A person may shift from compliance to adherence as motivation becomes internalized. However, the Hawthorne effect [39] can make this distinction unclear, as participants may appear adherent simply because they know they are being observed. In this study, the distribution of psychological themes across engagement clusters suggested more intrinsic motivation in the Consistent and Sporadic clusters and predominantly extrinsic motivation in Dropouts. These preliminary findings suggest that motivational profiles may influence engagement patterns and should be explored in larger and more diverse cohorts.

### 4.1 Limitations

The limited sample size (N=53) and high attrition rate (only 19% of participants used the app for more than 100 days) limit the generalizability and statistical power of the findings. Questionnaire response rates declined substantially over time (55% at T0, 30% at T1, 13% at T2), preventing meaningful longitudinal comparisons for most validated instruments. Additionally, 18 participants enrolled after September 2024 could not complete the full six-month study period. The number of patients invited to participate was not recorded, so the recruitment rate and the extent of selection bias cannot be quantified. Enrollment on a voluntary basis, with willingness to use digital technologies as an inclusion criterion, may have further biased the sample toward technologically literate and motivated individuals. Readiness for change, self-determination, and other psychological traits were not measured, limiting interpretation of motivational differences across the identified engagement clusters. Providing Fitbit wearables may also have inflated adherence, even though overall wearing time aligned with the engagement clusters. Differences in onboarding procedures between centers, such as the medication diary setup, were discovered only retrospectively and may have influenced usage patterns. The study was not registered in a clinical trial registry, so the distinction between pre-specified and exploratory analyses cannot be independently verified; all reported results should be interpreted as exploratory. Some questionnaires used were solution-specific and not validated. Non-random missingness could further amplify selection bias, requiring causal inference tools to properly address these sources of bias in future work. Moreover, statistical significance does not necessarily imply clinical relevance, underscoring the need for further research to derive meaningful clinical insights from the observational data.

### 4.2 Future work

Future work will incorporate higher-resolution wearable data to identify behavioral patterns and extend the analysis with latent class modeling using multi-modal signals. Subsequent research should explore digital biomarkers of engagement clusters, derive personalized insights into modifiable lifestyle factors, and evaluate the adaptive interventions using stratified or individualized models. In this study, we considered adaptive interventions solely for increasing engagement and did not evaluate them directly. Ethical restrictions prevented access to notification interaction data and information on potential background restrictions imposed by the operating system. Although findings from previous meta-analyses targeting physical activity through nudging reported only a small effect size (Cohen’s *d* = 0.31) [35], we plan to pursue stratified analyses and compare the main persuasive strategies to engagement clusters and psychological themes. Future design will emphasize user-centered customization, quick-response notifications, and adaptive personalization of the virtual companion through implicit user profiling with appropriate ethical safeguards [63].

### 4.3 Lessons learned for future design

Future development will emphasize user-centered and adaptive design. Users should be able to customize the home screen and prioritize relevant features. Quick-response notifications could streamline diary entries and enhance engagement. The Virtual Companion could incorporate adaptive personalization through implicit user profiling and psychological principles, as was employed for adaptive interventions in this study. With proper ethical safeguards, advances in artificial intelligence may enable more personalized, motivational, and diverse interactions. Finally, personalization of nudging algorithms can be further improved with personalized timing through just-in-time adaptive interventions and refined behavioral profiling [63], given sufficient real-time data granularity and scalability.

## Data Availability

The dataset analyzed during the current study is not publicly available to protect patient confidentiality.

## Acknowledgement

We thank Dr. Florent Krim and Evan Milstein from the Centre Hospitalier de Corbie, France, for their dedication to data collection and feedback, as well as Dr. Alexandre Antunes (Centro Hospitalar Leiria Pombal, Portugal), Dr. Giuseppe Santarpino (Città di Lecce Hospital, Italy), and András Gyöngy (Istituto EAHA di Prevenzione e Riabilitazione Cardiocircolatoria, Locarno e Valli, Switzerland). We also acknowledge the CUOREMA team at OSCAM (Fabrizio Pertile, Michele D’Andria, Marco Roncoroni, Emanuela Sala, Rosella Cozzi) and the Servizio di Cardi-ologia dell’Ospedale Malcantonese, Switzerland, for their participation. Last but not least, we thank Valerie van der Looij from Games for Health, and Alison Blythin and Dr. Adam Kirk from my mhealth Limited, for their contributions to the application development and study design.

## Data Availability Statement

The data underlying this article cannot be shared publicly due to ethical and data protection constraints. Anonymized summary data may be made available upon reasonable request to the corresponding author.

## Authors Contributions

R.Š.: Design and development of the nudging algorithm, Conceptualization of the study and analysis, Data curation, Methodology, Formal analysis, Writing - original draft, Correspondence.

D.M. and G.M.: Conceptualization of the study, Writing - review & editing. T.O.: Project funding, Conceptualization of the study, Data collection, Writing - review & editing. T.S.: Data collection. R.T.: Project funding, Conceptualization of the study, Design and deployment of Virtual Companion, deployment of the nudging algorithm, Writing - review & editing. R.L.: Project funding, Conceptualization of the study, Design of Virtual Companion, Writing - review & editing. N.B.: Design of Virtual Companion, Personal goals definition. W.R. and G.B.: Software. A.B.: Project administration. T.W.: Project funding. A.T.: Writing - review & editing. F.F.: Project funding, Conceptualization of the study, Writing - review & editing. All authors reviewed and approved the final version of the manuscript.

## Disclosure of Generative AI use

ChatGPT (OpenAI, GPT-5) was used to assist with language editing and refinement of wording. Claude (Anthropic, Fable 5) was used to assist with the preparation of graphical abstract. AI tools did not contribute to the study design, data analysis, or interpretation of results. All content was reviewed and approved by the authors.

## 5 Supplementary Materials

### 5.1 Full system specification

#### 5.1.1 Mobile application details

The mobile application includes the following sections:

##### Medication diary (MD)

Tracks medication adherence. Users select prescribed medications and schedules, then manually log each intake. Push notification reminders can be set at preferred times.

##### Activity diary (AD)

Users log daily steps and activities manually, or these are automati-cally retrieved from supported wearable devices. Manual entries include activity type, duration, distance, sessions, and rate of perceived exertion (RPE) on a 0–10 Borg scale [13; 72]. Auto-matically retrieved data contain daily step count, activity type, and duration.

##### Targets

Users set target ranges for physiological measures (steps, weight, BMI, blood pressure, glucose, cholesterol, LDL, HDL). A green/red indicator shows whether the latest value is within range.

##### Other self-monitoring sections

Additional sections support monitoring of fluid intake, weight, ECG images, echocardiograms, cholesterol, and other clinical exams.

##### Videos for education and exercises

Educational videos on heart physiology and conditions are provided. Exercise sessions include virtual reality-enhanced instructions and camera-based tracking. Walking videos simulating outdoor scenery can be followed at home. All sessions upload to the AD with self-reported RPE.

**Analytical platform** Summarizes patient data, such as medication intake and physical activity, helping clinicians make better-informed decisions in follow-up care.

### 5.2 Full inclusion criteria

- Be at least 18 years old and able to provide written informed consent
- Suitable to participate in an outpatient Phase II and Phase III CR program in telereha-bilitation
- Own a mobile device with Internet and Bluetooth connection, be able to constantly wear a smartwatch, and not have disabling motor limitations
- Be willing to use and interact with digital technologies, especially the CUOREMA solution running on mobile devices (phones and tablets) and with a smartwatch
- No unstable cardiovascular disease (e.g. accute myocardial infarction, coronary syndrome, …), no exclusion criteria for an outpatient CR and which currently require hospital admission, no disabling motor limitations, and no orthopedic and/or neurological condition that limits the ability to actively engage in training sessions

### 5.3 Solution in figures

### 5.4 Daily nudging algorithm

**Algorithm 1** Daily intervention selection and micro-randomization.

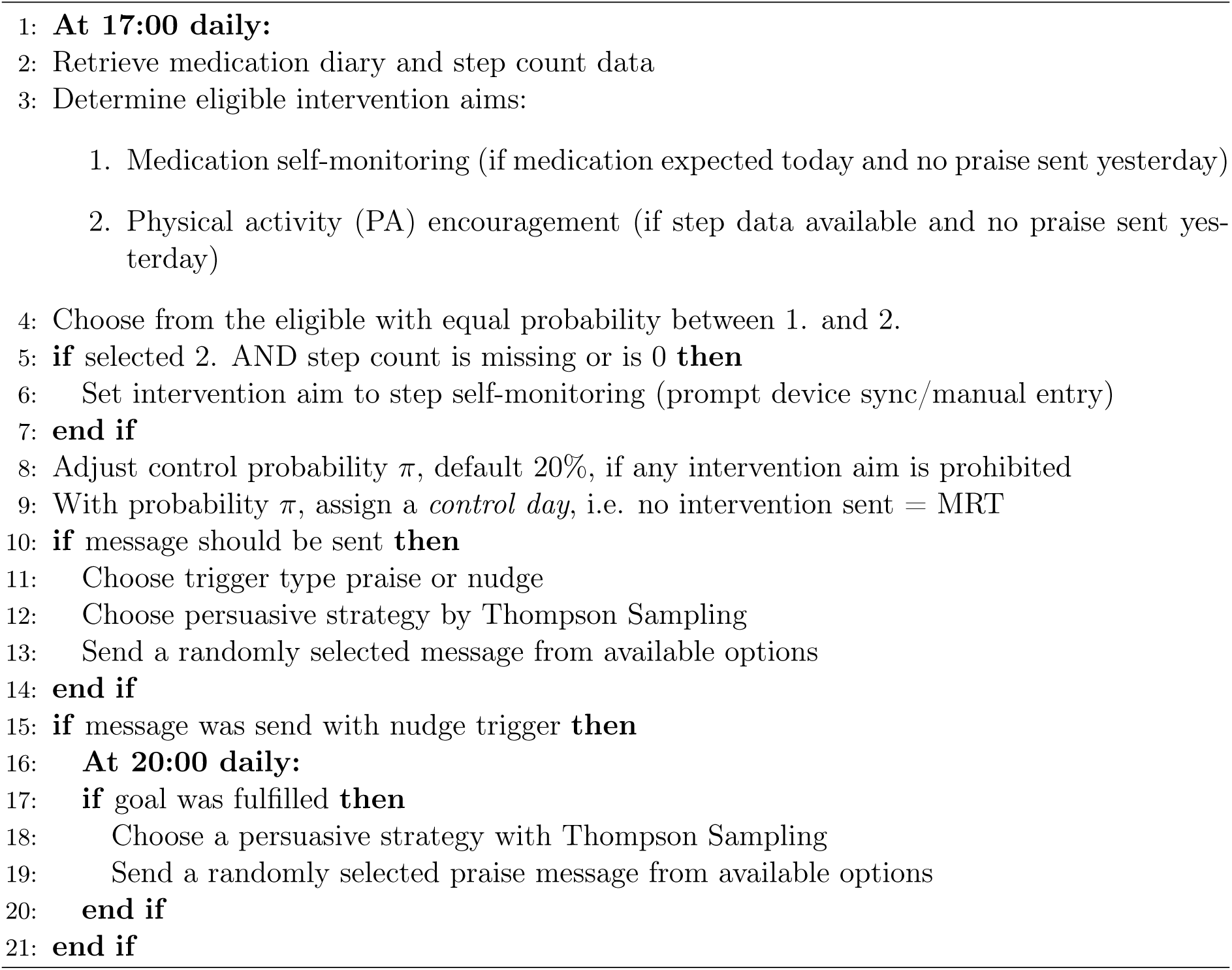

### 5.5 Study protocol schema

### 5.6 User experience and app usage results

**Figure 10:**
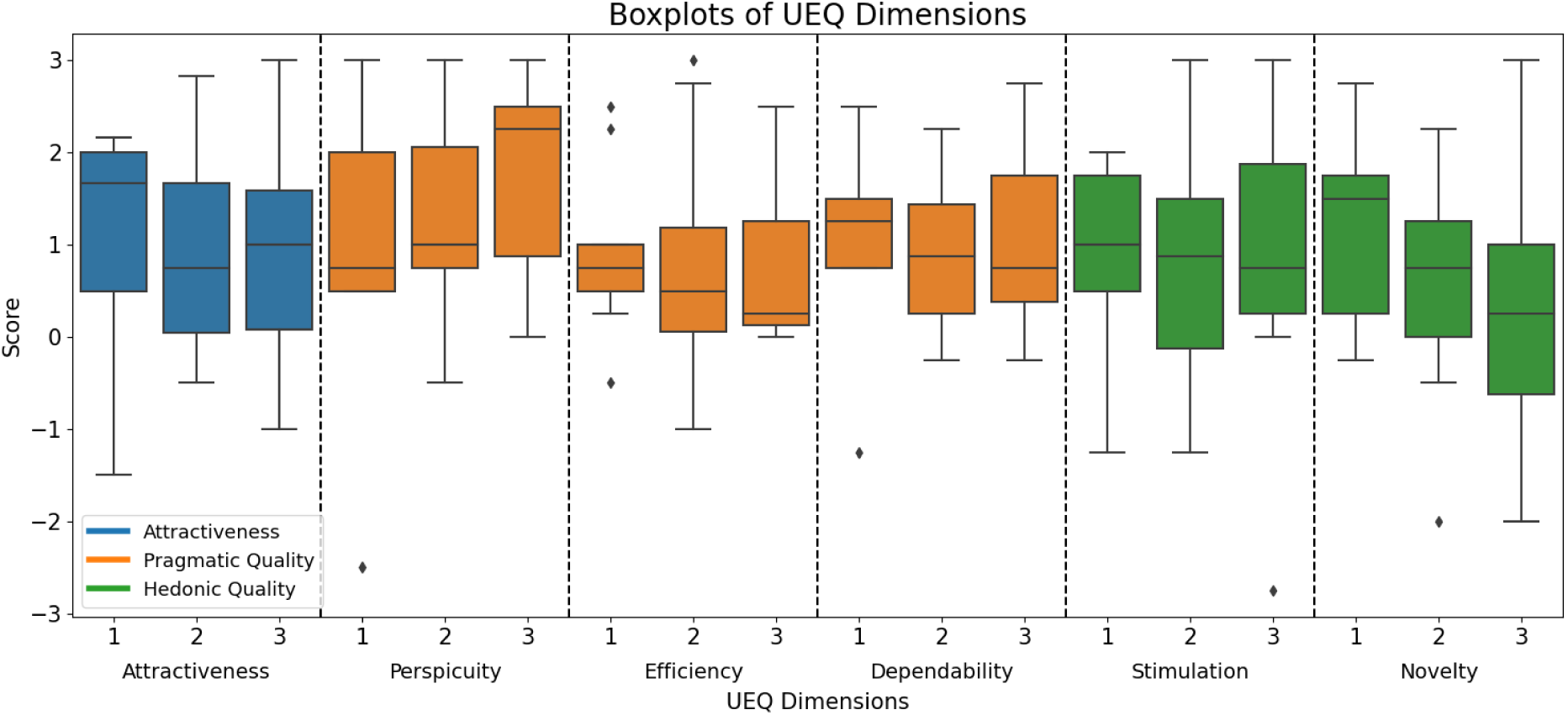
Results from UEQ questionnaire in 3 time-points, clustered to 3 quality types: attractiveness, pragmatic quality and hedonic quality.

**Table 5:** MD usage in the two rehabilitation centers.

|  | OSCAM | Corbie | p-value |
| --- | --- | --- | --- |
| Created medication therapy | 17 (94%) | 19 (54%) | **0.002** |
| MD used (active therapy) | 9 (53%) | 12 (63%) | 0.778 |
| Activated medication reminder | 7 (41%) | 2 (11%) | 0.204 |

**Table 6:** Distribution of user counts in number of interactions with virtual coaches, and median days to first and last interaction since T0.

| Virtual coach | N | N>1 | N≥10 | Days to<br>first inter-<br>action<br>(median days since T0) | Days to<br>last inter-<br>action |
| --- | --- | --- | --- | --- | --- |
| Cardiologist | 12 | 5 | 1 | 11.5 | 81 |
| Physiologist | 12 | 3 | 1 | 9.5 | 13.5 |
| Dietitian | 6 | 4 | 2 | 10.5 | 42 |
| Psychologist | 5 | 4 | 1 | 5 | 30 |
| Smoking cessation | 5 | 3 | 1 | 11 | 30 |

### 5.7 Solution-specific feedback questions

**Table 7:** N (%) of *positive* answers (mostly agree, agree, strongly agree) to questions tailored to the solution, distributed after the first month, at T1, and T2.

| Question | 1<sup>st</sup> mo.<br>(N=9) | T1<br>(N=13) | T2<br>(N=7) |
| --- | --- | --- | --- |
| **This programme helps me to...** |  |  |  |
| achieve my rehabilitation goals | 7 (78%) | 8 (61%) | 4 (57%) |
| improve my physical activity | 6 (67%) | 7 (53%) | 4 (57%) |
| improve my nutrition | 5 (56%) | 4 (30%) | 2 (28%) |
| improve my medication adherence | 2 (22%) | 3 (23%) | 3 (42%) |
| stop / reduce smoking | 4 (44%) | 2 (16%) | 3 (42%) |
| remember to monitor my vital parameters | 5 (56%) | 9 (69%) | 3 (42%) |
| learn more about my medical condition<br>and how to improve it | 5 (56%) | 8 (61%) | 3 (42%) |

**Table 8:** N (%) of *positive* answers (mostly agree, agree, strongly agree) to questions about motivational messages, distributed after the first month, at T1, and T2.

| Question | 1<sup>st</sup> mo.<br>(N=9) | T1<br>(N=13) | T2<br>(N=7) |
| --- | --- | --- | --- |
| Push notifications are delivered at<br>appropriate times | 5 (62%) | 2 (18%) | 1 (14%) |
| The content of the messages is motivational | 5 (62%) | 4 (33%) | 1 (14%) |
| **The motivational messages...** |  |  |  |
| are enjoyable and nice to read | 5 (62%) | 4 (33%) | 1 (14%) |
| are useful to help me remember to fill in<br>my medication diary | 2 (25%) | 3 (25%) | 3 (42%) |
| are useful to help me perform more<br>physical activity | 4 (50%) | 4 (33%) | 2 (28%) |
| are useful to help me remember to connect<br>my fitness tracker | 3 (37%) | 5 (41%) | 2 (28%) |

**Table 9:** N (%) of *positive* answers (mostly agree, agree, strongly agree) to questions about the Virtual Companion, distributed after the first month, at T1, and T2, asked only if participants reported using the Virtual Companion.

| Question | 1<sup>st</sup> mo.<br>(N=9) | T1<br>(N=13) | T2<br>(N=7) |
| --- | --- | --- | --- |
| The monthly review helps me to self-reflect | 4 (57%) | 3 (30%) | 2 (50%) |
| **The Virtual Companion...** |  |  |  |
| motivates me to use the system more than if it was not present at all | 5 (71%) | 1 (10%) | 2 (50%) |
| motivates me to stick to my rehabilitation goals and I would struggle otherwise | 5 (71%) | 2 (20%) | 2 (50%) |
| is a more enjoyable way to fill in my medication diary | 3 (42%) | 2 (20%) | 2 (50%) |
| helps me to better remember to fill in my medication diary | 1 (14%) | 1 (10%) | 3 (75%) |
| helps me to be more physically active | 5 (71%) | 1 (10%) | 2 (50%) |
| helps me to better remember to fill in my activity diary | 5 (71%) | 1 (10%) | 2 (50%) |
| is useful to help me learn useful things about how to improve my diet | 2 (28%) | 0 (0%) | 3 (75%) |
| is useful to help me relax | 3 (42%) | 0 (0%) | 2 (50%) |
| is useful on my journey to reduce or stop smoking | 2 (28%) | 0 (0%) | 2 (50%) |

**Table 10:** N (%) of *negative* answers (mostly disagree, disagree, strongly disagree) to solution-specific questions, distributed after the first month, at T1, and T2.

| **This program helps me to...** | 1<sup>st</sup> mo.<br>(N=9) | **T1**<br>(N=13) | **T2**<br>(N=7) |
| --- | --- | --- | --- |
| achieve my rehabilitation goals | 0 (0%) | 2 (15%) | 1 (14%) |
| improve my physical activity | 0 (0%) | 1 (7%) | 1 (14%) |
| improve my nutrition | 1 (11%) | 2 (15%) | 1 (14%) |
| improve my medication adherence | 1 (11%) | 1 (7%) | 1 (14%) |
| stop / reduce smoking | 2 (22%) | 1 (8%) | 2 (28%) |
| remember to monitor my vital parameters | 2 (22%) | 0 (0%) | 1 (14%) |
| learn more about my medical condition and how to improve it | 0 (0%) | 0 (0%) | 1 (14%) |

**Table 11:** N (%) of *negative* answers (mostly disagree, disagree, strongly disagree) to questions about motivational messages, distributed after the first month, at T1, and T2.

| Question | 1<sup>st</sup> mo.<br>(N=9) | T1<br>(N=13) | T2<br>(N=7) |
| --- | --- | --- | --- |
| Push notifications are delivered at appropriate times | 0 (0%) | 0 (0%) | 1 (14%) |
| The content of the messages is motivational | 0 (0%) | 4 (33%) | 2 (28%) |
| **The motivational messages...** |  |  |  |
| are enjoyable and nice to read | 1 (12%) | 4 (33%) | 2 (28%) |
| are useful to help me remember to fill in my medication diary | 4 (50%) | 2 (16%) | 1 (14%) |
| are useful to help me perform more physical activity | 1 (12%) | 3 (25%) | 1 (14%) |
| are useful to help me remember to connect my fitness tracker | 2 (25%) | 3 (25%) | 2 (28%) |

**Table 12:** N (%) of *negative* answers (mostly disagree, disagree, strongly disagree) to questions about Virtual Companion, distributed after 1st month, at T1 and T2, asked if participants reported using Virtual Companion.

| Question | 1<sup>st</sup> mo.<br>(N=9) | T1<br>(N=13) | T2<br>(N=7) |
| --- | --- | --- | --- |
| The monthly review helps me to self-reflect | 1 (14%) | 1 (10%) | 1 (25%) |
| **The virtual companion...** |  |  |  |
| motivates me to use the system more than if it was not present at all | 0 (0%) | 4 (40%) | 2 (50%) |
| motivates me to stick to my rehabilitation goals and I would struggle otherwise | 1 (14%) | 2 (20%) | 1 (25%) |
| is more a enjoyable way to fill in my medication diary | 3 (42%) | 2 (20%) | 2 (50%) |
| helps me to better remember to fill in my medication diary | 2 (28%) | 0 (0%) | 1 (25%) |
| helps me to be more physically active | 1 (14%) | 2 (20%) | 1 (25%) |
| helps me to better remember to fill in my activity diary | 0 (0%) | 1 (10%) | 1 (25%) |
| is useful to help me learn useful things about how to improve my diet | 1 (14%) | 1 (10%) | 1 (25%) |
| is useful to help me relax | 1 (14%) | 3 (30%) | 1 (25%) |
| is useful on my journey to reduce or stop smoking | 2 (28%) | 1 (11%) | 2 (50%) |

### 5.8 Summary statistics of health-related questionnaires and clinical test 6MWT

**Table 13:** Results of paired one-sided Wilcoxon tests on the improvements between T0 and T1 in the health-related validated questionnaires.

| Questionnaire | Sample size | N improved (the same) | p-value |
| --- | --- | --- | --- |
| GAD | 12 | 4 (6) | 0.312 |
| PHQ | 12 | 6 (3) | 0.275 |
| Heart-QoL | 13 | 9 (1) | 0.025 |
| Morisky scale | 12 | 4 (6) | 0.125 |

**Table 14:** Comparison of questionnaires Scores between Sporadic and Consistent engagement clusters at T0. Reported are median [*p*_25_*, p*_75_] and p-values from Mann-Whitney U test.

|  | Sporadic | Consistent | p-value |
| --- | --- | --- | --- |
| PHQ | 2.00 [1.00, 4.00] | 2.00 [2.00, 7.00] | 0.362 |
| GAD | 3.00 [0.00, 5.00] | 3.00 [1.50, 5.50] | 0.703 |
| QoL Emotional | 2.50 [2.50, 3.00] | 2.25 [1.62, 2.62] | 0.124 |
| QoL Physical | 2.40 [2.20, 2.70] | 1.90 [1.15, 2.45] | 0.072 |
| QoL Total | 2.50 [2.43, 2.64] | 2.21 [1.32, 2.36] | 0.051 |
| Morisky scale | 8.00 [7.00, 8.00] | 8.00 [8.00, 8.00] | 0.200 |

**Table 15:** Statistics of GAD questionnaire scores in the 3 time points. Lower score is better. Maximal score in GAD is 21 [60]. Values of 4 or less mean no to minimal anxiety, and 5-9 or less mild anxiety.

| | N | Min | Max | Mean | Median | Std | Score $\leq 4$ | Score 5-9 |
| --- | --- | --- | --- | --- | --- | --- | --- | --- |
| T0 | 28 | 0 | 19 | 4 | 4 | 4 | 16 (57%) | 10 (36%) |
| T1 | 15 | 0 | 7 | 2 | 1 | 3 | 11 (73%) | 4 (27%) |
| T2 | 7 | 0 | 6 | 2 | 2 | 2 | 5 (71%) | 2 (29%) |

**Table 16:** Statistics of PHQ questionnaire scores in the 3 timepoints. Lower score is better. Maximal value of PHQ is 27 [32]. The score of 4 or below signifies no to minimal depression, whereas score of 5-9 means mild depression.

| | N | Min | Max | Mean | Median | Std | Score $\leq 4$ | Score 5-9 |
| --- | --- | --- | --- | --- | --- | --- | --- | --- |
| T0 | 28 | 0 | 22 | 5 | 2 | 5 | 18 (64%) | 7 (25%) |
| T1 | 15 | 0 | 9 | 3 | 2 | 3 | 11 (73%) | 4 (27%) |
| T2 | 7 | 0 | 8 | 3 | 2 | 3 | 5 (71%) | 2 (29%) |

**Table 17:** Statistics of the emotional, physical components, and total score of Heart-QoL ques-tionnaire scores in the 3 time points. Higher score is better. The maximum possible score of Heart-QoL is 3. Emotional and Physical components are averaged and the total score is ob-tained.

| Component | | N | Min | Max | Mean | Median | Std | Score $\leq 1$ |
| --- | --- | --- | --- | --- | --- | --- | --- | --- |
| Emotional | T0 | 28 | 0.0 | 3.0 | 2.2 | 2.5 | 0.8 | 3 (11%) |
| Emotional | T1 | 16 | 1.8 | 3.0 | 2.6 | 2.5 | 0.4 | 0 (0%) |
| Emotional | T2 | 7 | 1.8 | 3.0 | 2.6 | 2.8 | 0.5 | 0 (0%) |
| Physical | T0 | 28 | 0.2 | 2.9 | 2.0 | 2.2 | 0.8 | 5 (18%) |
| Physical | T1 | 16 | 0.0 | 3.0 | 2.3 | 2.8 | 0.9 | 1 (6%) |
| Physical | T2 | 7 | 0.9 | 3.0 | 2.6 | 2.8 | 0.7 | 1 (14%) |
| Total | T0 | 28 | 0.2 | 2.8 | 2.0 | 2.3 | 0.7 | 4 (14%) |
| Total | T1 | 16 | 0.7 | 3.0 | 2.4 | 2.8 | 0.7 | 1 (6%) |
| Total | T2 | 7 | 1.1 | 3.0 | 2.6 | 2.8 | 0.7 | 0 (0%) |

**Table 18:** Statistics of Morisky medication adherence questionnaire scores in the 3 time points. Higher score is better. Maximal value of Morisky scale [11; 15] is 8, and score 7 is still considered as adherent.

|  | N | Min | Max | Mean | Median | Std |
| --- | --- | --- | --- | --- | --- | --- |
| T0 | 28 | 5 | 8 | 7 | 8 | 1 |
| T1 | 15 | 7 | 8 | 8 | 8 | 0 |
| T2 | 7 | 6 | 8 | 8 | 8 | 1 |

**Table 19:** Summary statistics of 6MWT tests of all measured participants.

| | N | Mean | Std | Median [$p_{25}$, $p_{75}$] |
| --- | --- | --- | --- | --- |
| T0 | 19 | 511.32 | 81.29 | 527.00 [479.00, 554.50] |
| T1 | 14 | 599.14 | 94.41 | 604.50 [587.00, 658.00] |
| T2 | 11 | 608.64 | 143.87 | 603.00 [547.50, 659.50] |

## Footnotes

1 https://mymhealth.com/myheart

## References

[1] Alan Agresti and Maria Kateri. Categorical data analysis. In International encyclopedia of statistical science, pages 206–208. Springer, 2011.

[2] Leonardo Angelini, Mira El Kamali, Elena Mugellini, Omar Abou Khaled, Christina Röcke, Simone Porcelli, Alfonso Mastropietro, Giovanna Rizzo, Noemi Boqué, Josep Maria Del Bas, et al. The nestore e-coach: designing a multi-domain pathway to well-being in older age. Technologies, 10(2):50, 2022.

[3] Victoria Arija, Felipe Villalobos, Roser Pedret, Angels Vinuesa, Dolors Jovani, Gabriel Pascual, and Josep Basora. Physical activity, cardiovascular health, quality of life and blood pressure control in hypertensive subjects: randomized clinical trial. Health and quality of life outcomes, 16(1):184, 2018.

[4] Y Arima, T Kaihara, S Takizawa, S Doi, K Honda, Y Kobayashi, A Kasagawa, Y Kawagoe, T Suzuki, T Kai, et al. Home-based cardiac rehabilitation with a smartwatch and a smart-phone gamification application: a pilot study. European Journal of Preventive Cardiology, 31(Supplement_1):zwae175–119, 2024.

[5] Authors/Task Force Members, Massimo F Piepoli, Arno W Hoes, Stefan Agewall, Christian Albus, Carlos Brotons, Alberico L Catapano, Marie-Therese Cooney, Ugo Corrà, Bernard Cosyns, et al. 2016 european guidelines on cardiovascular disease prevention in clinical practice: The sixth joint task force of the european society of cardiology and other societies on cardiovascular disease prevention in clinical practice (constituted by representatives of 10 societies and by invited experts): Developed with the special contribution of the european association for cardiovascular prevention & rehabilitation (eacpr). European journal of preventive cardiology, 23(11):NP1–NP96, 2016.

[6] Claudia Avina. The use of self-monitoring as a treatment intervention. In Evidence-based adjunctive treatments, pages 207–219. Elsevier, 2008.

[7] Ladislav Batalik, Katerina Filakova, Michaela Sladeckova, Filip Dosbaba, Jingjing Su, and Garyfallia Pepera. The cost-effectiveness of exercise-based cardiac telerehabilitation intervention: a systematic review. EuropEan Journal of physical and rEhabilitation MEdicinE, 59(2):248, 2023.

[8] Hannes Baumann, Ben Singh, Staiano E Amanda, Claire Gough, Mavra Ahmed, Janis Fiedler, Kathrin Wunsch, Alyssa Button, Zenong Yin, Maria Vasiloglou, et al. Effectiveness of mhealth interventions targeting physical activity, sedentary behaviour, sleep or nutrition on emotional, behavioural and eating disorders in adolescents: A systematic review and meta-analysis. Frontiers in Digital Health, 7:1593677.

[9] R Nicole Bellet, Lewis Adams, and Norman R Morris. The 6-minute walk test in outpatient cardiac rehabilitation: validity, reliability and responsiveness—a systematic review. Physiotherapy, 98(4):277–286, 2012.

[10] Jacob Benesty, Jingdong Chen, Yiteng Huang, and Israel Cohen. Pearson correlation coefficient. In Noise reduction in speech processing, pages 1–4. Springer, 2009.

[11] Dan R Berlowitz, Capri G Foy, Lewis E Kazis, Linda P Bolin, Molly B Conroy, Peter Fitzpatrick, Tanya R Gure, Paul L Kimmel, Kent Kirchner, Donald E Morisky, et al. Effect of intensive blood-pressure treatment on patient-reported outcomes. New England Journal of Medicine, 377(8):733–744, 2017.

[12] Carlo Bonferroni. Teoria statistica delle classi e calcolo delle probabilita. Pubblicazioni del R istituto superiore di scienze economiche e commericiali di firenze, 8:3–62, 1936.

[13] Gunnar A Borg. Psychophysical bases of perceived exertion. Medicine and science in sports and exercise, 14(5):377–381, 1982.

[14] Ronald D Boschloo. Raised conditional level of significance for the 2*×* 2-table when testing the equality of two probabilities. Statistica Neerlandica, 24(1):1–9, 1970.

[15] Adam P Bress, Brandon K Bellows, Jordan B King, Rachel Hess, Srinivasan Beddhu, Zugui Zhang, Dan R Berlowitz, Molly B Conroy, Larry Fine, Suzanne Oparil, et al. Cost-effectiveness of intensive versus standard blood-pressure control. New England Journal of Medicine, 377(8):745–755, 2017.

[16] Eric R Buhi, Tara E Trudnak, Mary P Martinasek, Alison B Oberne, Hollie J Fuhrmann, and Robert J McDermott. Mobile phone-based behavioural interventions for health: A systematic review. Health Education Journal, 72(5):564–583, 2013.

[17] Maurizio Caon, Federica Prinelli, Leonardo Angelini, Stefano Carrino, Elena Mugellini, Silvia Orte, José CE Serrano, Sarah Atkinson, Anne Martin, and Fulvio Adorni. Pegaso e-diary: User engagement and dietary behavior change of a mobile food record for adolescents. Frontiers in Nutrition, 9:727480, 2022.

[18] Robert Cialdini. Influence: Science and Practice. 01 1993.

19. Cameron Davidson-Pilon. lifelines: survival analysis in python. Journal of Open Source Software, 4(40):1317, 2019. doi: 10.21105/joss.01317. URL 10.21105/joss.01317.

[20] Shannon M Dunlay, Brandi J Witt, Thomas G Allison, Sharonne N Hayes, Susan A Weston, Ellen Koepsell, and Véronique L Roger. Barriers to participation in cardiac rehabilitation. American heart journal, 158(5):852–859, 2009.

[21] Bambang Dwiputra, Anwar Santoso, Budhi Setianto Purwowiyoto, Basuni Radi, Ade Meidian Ambari, Dwita Rian Desandri, Serlie Fatrin, and Bashar Adi Wahyu Pandhita. Smartphone-based cardiac rehabilitation program improves functional capacity in coronary heart disease patients: a systematic review and meta-analysis. Global heart, 18(1):42, 2023.

[22] Ronald A Fisher. On the interpretation of *χ* 2 from contingency tables, and the calculation of p. Journal of the royal statistical society, 85(1):87–94, 1922.

23. Brian J Fogg. Tiny habits: The small changes that change everything. Harvest, 2020.

[24] Benjamin Gardner, Phillippa Lally, and Jane Wardle. Making health habitual: the psychology of ‘habit-formation’and general practice. The British Journal of General Practice, 62(605):664, 2012.

25. Myles Hollander, Douglas A Wolfe, and Eric Chicken. Nonparametric statistical methods. John Wiley & Sons, 2013.

[26] FN Huq, NAM Momenuzzaman, AW Chowdhury, MM Hoque, KN Khan, F Begum, AM Shafique, R Anis, MA Rahman, S Nahar, et al. Effect of telephone-monitored home-based cardiac rehabilitation exercise on functional capacity and quality of life in heart failure patients in a lower-middle-income country. European Journal of Preventive Cardiology, 29 (Supplement_1):zwac056–248, 2022.

[27] Robert Jakob, Samira Harperink, Aaron Maria Rudolf, Elgar Fleisch, Severin Haug, Jacqueline Louise Mair, Alicia Salamanca-Sanabria, and Tobias Kowatsch. Factors influencing adherence to mhealth apps for prevention or management of noncommunicable diseases: systematic review. Journal of medical Internet research, 24(5):e35371, 2022.

[28] Nan Jiang, Yongfa Wu, and Chengjia Li. Limitations of using cox proportional hazards model in cardiovascular research. Cardiovascular Diabetology, 23(1):219, 2024.

[29] Maurits Kaptein, Panos Markopoulos, Boris de Ruyter, and Emile Aarts. Personalizing persuasive technologies: Explicit and implicit personalization using persuasion profiles. International Journal of Human-Computer Studies, 77:38–51, 2015. ISSN 1071-5819. doi: 10.1016/j.ijhcs.2015.01.004.

[30] Predrag Klasnja, Eric B Hekler, Saul Shiffman, Audrey Boruvka, Daniel Almirall, Ambuj Tewari, and Susan A Murphy. Microrandomized trials: An experimental design for developing just-in-time adaptive interventions. Health Psychology, 34(S):1220, 2015.

[31] Kornelia Kotseva, David Wood, Dirk De Bacquer, Guy De Backer, Lars Rydén, Catriona Jennings, Viveca Gyberg, Philippe Amouyel, Jan Bruthans, Almudena Castro Conde, et al. Euroaspire iv: A european society of cardiology survey on the lifestyle, risk factor and therapeutic management of coronary patients from 24 european countries. European journal of preventive cardiology, 23(6):636–648, 2016.

[32] Kurt Kroenke, Robert L Spitzer, and Janet BW Williams. The phq-9: validity of a brief depression severity measure. Journal of general internal medicine, 16(9):606–613, 2001.

[33] William H Kruskal and W Allen Wallis. Use of ranks in one-criterion variance analysis. Journal of the American statistical Association, 47(260):583–621, 1952.

[34] Stuart Lloyd. Least squares quantization in pcm. IEEE transactions on information theory, 28(2):129–137, 1982.

[35] Haoming Ma, Aoqi Wang, Runyuan Pei, and Meihua Piao. Effects of habit formation interventions on physical activity habit strength: meta-analysis and meta-regression. International Journal of Behavioral Nutrition and Physical Activity, 20(1):109, 2023.

[36] Ralph Maddison, Jonathan Charles Rawstorn, Ralph AH Stewart, Jocelyne Benatar, Robyn Whittaker, Anna Rolleston, Yannan Jiang, Lan Gao, Marj Moodie, Ian Warren, et al. Effects and costs of real-time cardiac telerehabilitation: randomised controlled non-inferiority trial. Heart, 105(2):122–129, 2019.

[37] Mohsen Masoumian Hosseini, Seyedeh Toktam Masoumian Hosseini, Karim Qayumi, Shahriar Hosseinzadeh, and Seyedeh Saba Sajadi Tabar. Smartwatches in healthcare medicine: assistance and monitoring; a scoping review. BMC Medical Informatics and Decision Making, 23(1):248, 2023.

[38] Theodoros Maximidou and Ute Mons. Benefits of wearable-based cardiac rehabilitation interventions in secondary prevention of coronary artery disease–a systematic review and meta-analysis. American Journal of Preventive Cardiology, page 101015, 2025.

[39] Rob McCarney, James Warner, Steve Iliffe, Robbert Van Haselen, Mark Griffin, and Peter Fisher. The hawthorne effect: a randomised, controlled trial. BMC medical research methodology, 7(1):30, 2007.

[40] Sinead TJ McDonagh, Hasnain Dalal, Sarah Moore, Christopher E Clark, Sarah G Dean, Kate Jolly, Aynsley Cowie, Jannat Afzal, and Rod S Taylor. Home-based versus centre-based cardiac rehabilitation. Cochrane database of systematic reviews, (10), 2023.

[41] Tasaduq H Mir. Adherence versus compliance. HCA healthcare journal of medicine, 4(2): 219, 2023.

[42] John Ashworth Nelder and Robert WM Wedderburn. Generalized linear models. Journal of the Royal Statistical Society Series A: Statistics in Society, 135(3):370–384, 1972.

[43] Ryan Ng, Rinku Sutradhar, Zhan Yao, Walter P Wodchis, and Laura C Rosella. Smoking, drinking, diet and physical activity—modifiable lifestyle risk factors and their associations with age to first chronic disease. International journal of epidemiology, 49(1):113–130, 2020.

[44] Neil Oldridge, Stefan Höfer, Hannah McGee, Ronan Conroy, Frank Doyle, Hugo Saner, and HeartQoL Project Investigators). The heartqol: Part i. development of a new core health-related quality of life questionnaire for patients with ischemic heart disease. European journal of preventive cardiology, 21(1):90–97, 2014.

[45] Kristina Orth-Gomér, Annika Rosengren, and Lars Wilhelmsen. Lack of social support and incidence of coronary heart disease in middle-aged swedish men. Biopsychosocial Science and Medicine, 55(1):37–43, 1993.

[46] Nicholas Ownbey, Jeff Soukup, Elizabeth Fugate-Whitlock, and Tina MK Newsham. Evaluation of telephone-based cardiac rehabilitation services delivered to adults 65 and older during the early months of the covid-19 pandemic. Journal of Applied Gerontology, 41(10): 2226–2234, 2022.

[47] F. Pedregosa, G. Varoquaux, A. Gramfort, V. Michel, B. Thirion, O. Grisel, M. Blondel, P. Prettenhofer, R. Weiss, V. Dubourg, J. Vanderplas, A. Passos, D. Cournapeau, M. Brucher, M. Perrot, and E. Duchesnay. Scikit-learn: Machine learning in Python. Journal of Machine Learning Research, 12:2825–2830, 2011.

[48] Antonio Pelliccia, Sanjay Sharma, Sabiha Gati, Maria Bäck, Mats Börjesson, Stefano Caselli, Jean-Philippe Collet, Domenico Corrado, Jonathan A Drezner, Martin Halle, et al. 2020 esc guidelines on sports cardiology and exercise in patients with cardiovascular disease: The task force on sports cardiology and exercise in patients with cardiovascular disease of the european society of cardiology (esc). European heart journal, 42(1):17–96, 2021.

[49] Lukasz Piwek, David A Ellis, Sally Andrews, and Adam Joinson. The rise of consumer health wearables: promises and barriers. PLoS medicine, 13(2):e1001953, 2016.

[50] James O Prochaska and Wayne F Velicer. The transtheoretical model of health behavior change. American journal of health promotion, 12(1):38–48, 1997.

[51] Ulrich Reimer, Edith Maier, and Tom Ulmer. A self-learning application framework for behavioral change support. In Information and Communication Technologies for Ageing Well and e-Health: Second International Conference, ICT4AWE 2016, Rome, Italy, April 21-22, 2016, Revised Selected Papers 2, pages 119–139. Springer, 2017.

[52] Davinia Maria Resurrección, Patricia Moreno-Peral, Marta Gomez-Herranz, Maria Rubio-Valera, Luis Pastor, Jose Miguel Caldas de Almeida, and Emma Motrico. Factors associated with non-participation in and dropout from cardiac rehabilitation programmes: a systematic review of prospective cohort studies. European Journal of Cardiovascular Nursing, 18 (1):38–47, 2019.

[53] Daniel J Russo, Benjamin Van Roy, Abbas Kazerouni, Ian Osband, Zheng Wen, et al. A tutorial on thompson sampling. Foundations and Trends® in Machine Learning, 11(1): 1–96, 2018.

[54] Richard M Ryan and Edward L Deci. Self-determination theory and the facilitation of intrinsic motivation, social development, and well-being. American psychologist, 55(1):68, 2000.

[55] Richard M Ryan and Edward L Deci. Intrinsic and extrinsic motivation from a self-determination theory perspective: Definitions, theory, practices, and future directions. Contemporary educational psychology, 61:101860, 2020.

[56] Martijn Scherrenberg, Maarten Falter, Ana Abreu, Suleman Aktaa, Stefan Busnatu, Ruben Casado-Arroyo, Paul Dendale, Polychronis Dilaveris, Emanuela T Locati, Elena Marques-Sule, et al. Standards for cardiac telerehabilitation: A scientific statement of the european association of preventive cardiology (eapc) and the association of cardiovascular nursing & allied professions (acnap) of the esc, and the esc working group on e-cardiology. European Heart Journal, page ehaf408, 2025.

[57] Patrick Schober and Thomas R Vetter. Survival analysis and interpretation of time-to-event data: the tortoise and the hare. Anesthesia & Analgesia, 127(3):792–798, 2018.

[58] Martin Schrepp. User experience questionnaire handbook. All you need to know to apply the UEQ successfully in your project, 10, 2015.

[59] Wejdan Shahin, Gerard A Kennedy, and Ieva Stupans. The association between social support and medication adherence in patients with hypertension: A systematic review. Pharmacy Practice (Granada), 19(2), 2021.

[60] Robert L Spitzer, Kurt Kroenke, Janet BW Williams, and Bernd Löwe. A brief measure for assessing generalized anxiety disorder: the gad-7. Archives of internal medicine, 166 (10):1092–1097, 2006.

[61] Sabine Stamm-Balderjahn, Sebastian Bernert, and Susanne Rossek. Promoting patient self-management following cardiac rehabilitation using a web-based application: a pilot study. Digital Health, 9:20552076231211546, 2023.

[62] Madoka Sunamura, Nienke Ter Hoeve, Rita JG van den Berg-Emons, Eric Boersma, Marcel L Geleijnse, and Ron T van Domburg. Patients who do not complete cardiac rehabilitation have an increased risk of cardiovascular events during long-term follow-up. Netherlands Heart Journal, 28(9):460–466, 2020.

[63] Radoslava Švihrová, Alvise Dei Rossi, Davide Marzorati, Athina Tzovara, and Francesca Dalia Faraci. Designing digital health interventions with causal inference and multi-armed bandits: A review. Frontiers in Digital Health, 7, 2025.

[64] Rod S Taylor, Hayes Dalal, Kate Jolly, Tiffany Moxham, and Anna Zawada. Home-based versus centre-based cardiac rehabilitation. Cochrane database of systematic reviews, (1), 2010.

65. Joseph Tessler, Intisar Ahmed, and Bruno Bordoni. Cardiac rehabilitation. In StatPearls [Internet]. StatPearls Publishing, 2025.

[66] Francesco Trovo, Stefano Paladino, Marcello Restelli, and Nicola Gatti. Sliding-window thompson sampling for non-stationary settings. Journal of Artificial Intelligence Research, 68:311–364, 2020.

[67] Pauli Virtanen, Ralf Gommers, Travis E. Oliphant, Matt Haberland, Tyler Reddy, David Cournapeau, Evgeni Burovski, Pearu Peterson, Warren Weckesser, Jonathan Bright, Stéfan J. van der Walt, Matthew Brett, Joshua Wilson, K. Jarrod Millman, Nikolay Mayorov, Andrew R. J. Nelson, Eric Jones, Robert Kern, Eric Larson, C J Carey, İlhan Polat, Yu Feng, Eric W. Moore, Jake VanderPlas, Denis Laxalde, Josef Perktold, Robert Cimrman, Ian Henriksen, E. A. Quintero, Charles R. Harris, Anne M. Archibald, Antônio H. Ribeiro, Fabian Pedregosa, Paul van Mulbregt, and SciPy 1.0 Contributors. SciPy 1.0: Fundamental Algorithms for Scientific Computing in Python. Nature Methods, 17:261–272, 2020. doi: 10.1038/s41592-019-0686-2.

[68] Frank LJ Visseren, François Mach, Yvo M Smulders, David Carballo, Konstantinos C Koskinas, Maria Bäck, Athanase Benetos, Alessandro Biffi, José-Manuel Boavida, Davide Capodanno, et al. 2021 esc guidelines on cardiovascular disease prevention in clinical practice: Developed by the task force for cardiovascular disease prevention in clinical practice with representatives of the european society of cardiology and 12 medical societies with the special contribution of the european association of preventive cardiology (eapc). European heart journal, 42(34):3227–3337, 2021.

[69] Christiaan Vrints, Felicita Andreotti, Konstantinos C Koskinas, Xavier Rossello, Marianna Adamo, James Ainslie, Adrian Paul Banning, Andrzej Budaj, Ronny R Buechel, Giovanni Alfonso Chiariello, et al. 2024 esc guidelines for the management of chronic coronary syndromes: developed by the task force for the management of chronic coronary syndromes of the european society of cardiology (esc) endorsed by the european association for cardio-thoracic surgery (eacts). European heart journal, 45(36):3415–3537, 2024.

[70] Fan Wang, Yu Gao, Zhen Han, Yue Yu, Zhiping Long, Xianchen Jiang, Yi Wu, Bing Pei, Yukun Cao, Jingyu Ye, et al. A systematic review and meta-analysis of 90 cohort studies of social isolation, loneliness and mortality. Nature human behaviour, 7(8):1307–1319, 2023.

71. R Jay Widmer, Nerissa M Collins, C Scott Collins, Colin P West, Lilach O Lerman, and Amir Lerman. Digital health interventions for the prevention of cardiovascular disease: a systematic review and meta-analysis. In Mayo Clinic Proceedings, volume 90, pages 469–480. Elsevier, 2015.

[72] Nerys Williams. The borg rating of perceived exertion (rpe) scale. Occupational medicine, 67(5):404–405, 2017.

[73] World Medical Association. World Medical Association Declaration of Helsinki: Ethical principles for medical research involving human subjects. JAMA, 310(20):2191–2194, November 2013. ISSN 1538-3598. doi: 10.1001/jama.2013.281053.

[74] Ashkan Dehghani Zahedani, Tracey McLaughlin, Arvind Veluvali, Nima Aghaeepour, Amir Hosseinian, Saransh Agarwal, Jingyi Ruan, Shital Tripathi, Mark Woodward, Noosheen Hashemi, et al. Digital health application integrating wearable data and behavioral patterns improves metabolic health. NPJ digital medicine, 6(1):216, 2023.

